# Proteomic resolution of the MASLD cardiometabolic spectrum identifies sex-driven endotypes and predicts systemic mortality

**DOI:** 10.64898/2025.12.02.25341467

**Authors:** Luis Diambra, Silvia Sookoian, Carlos Jose Pirola

## Abstract

**Background:** Metabolic dysfunction-associated steatotic liver disease (MASLD) represents a systemic metabolic hub where overlapping cardiometabolic risk factors frequently obscure individualized prognostic trajectories. While MASLD is a known driver of systemic risk, the molecular endotypes that dictate clinical outcomes across the disease spectrum remain poorly defined. We utilized a population-wide proteomic framework to resolve the MASLD cardiometabolic spectrum, identifying the sex-driven molecular endotypes that bridge hepatic dysfunction with systemic mortality.

**Methods:** We applied latent class analysis (LCA) to 48,806 UK Biobank participants with MRI-PDFF data to define phenotypic clusters across a >15-year follow-up. Large-scale proteomic profiling was integrated to characterize the molecular architecture of these identified clusters. This led to the derivation of a four-protein score (PS4), which was robustly validated for all-cause mortality prediction in an expanded cohort (n=115,979) and an external replication cohort. Causal mediation analysis was employed to quantify the biological contribution of these circulating and hepatic-expressed proteins to the observed clinical outcomes.

**Results:** The LCA resolved the MASLD spectrum into clinically distinct endotypes, revealing a pronounced sexual dimorphism where high-risk subgroups displayed accelerated cardiometabolic decline. The PS4 score—comprising proteins linked to hepatic lipid metabolism (FABP1), one-carbon metabolism (FTCD), and immune-vascular integrity (ADGRG1, GAST)—demonstrated superior performance in predicting systemic mortality compared to established clinical scores. Mediation analysis revealed that these proteins account for a substantial causal fraction (10–24%) of the association between MASLD endotypes and mortality, independent of traditional risk factors such as age, sex, hypertension and type 2 diabetes.

**Conclusions:** Our findings demonstrate that MASLD-driven proteomic shifts provide a mechanistic link between hepatic metabolic dysfunction and systemic survival. By resolving the MASLD cardiometabolic spectrum, we identified high-risk endotypes that are not captured by traditional liver-centric scores. The PS4 signature represents a biologically grounded framework for deciphering the systemic consequences of MASLD and its associated cardiometabolic complications, offering a scalable tool to identify individuals at the highest risk for adverse outcomes and facilitating a precision medicine approach to cardiometabolic disease.

## INTRODUCTION

Metabolic dysfunction-associated steatotic liver disease (MASLD), recently updated from non-alcoholic fatty liver disease (NAFLD), is now recognized as the most prevalent chronic liver condition worldwide in both children and adults [1]. The current disease definition involves the presence of steatosis with at least one of five cardiometabolic risk factors [2]. The MASLD clinical spectrum is rooted in identifiable histopathological stages, spanning from relatively benign steatosis to the inflammatory, fibrotic condition known as metabolic dysfunction-associated steatohepatitis (MASH) [1,3]. The progression of MASH to severe conditions, such as cirrhosis and hepatocellular carcinoma (HCC), is associated with high morbidity and mortality [4] [5]. In parallel, MASLD is a multi-systemic disorder associated with increased all-cause mortality and a heightened risk for nearly all major extra-hepatic comorbidities, including cardiovascular disease (CVD) [6] and non-HCC cancer [7].

Recent studies underscore a crucial aspect of the disease biology: substantial disease heterogeneity [8–10]. This term describes the existence of diverse, often hidden, sub-phenotypes that manifest in individuals despite similar diagnostic criteria. This complexity stems from the combined and overlapping heterogeneity across the underlying risk factors and comorbidities captured within the new disease definition, thus contributing to the challenge of predicting individual patient outcomes. Specifically, the diseases comprising the metabolic syndrome (MetS), now integral to the MASLD definition—including type 2 diabetes (T2DM), obesity, dyslipidaemia, and arterial hypertension—are themselves characterized by diverse pathophysiological processes and heterogeneous risk profiles [11,12]. Importantly, MASLD is established as a principal component of the MetS, often sharing underlying biological and genetic mechanisms with these co-occurring conditions [13].

To dissect this complexity, studies have leveraged data-driven clustering analyses to delineate heterogeneity [14] and applied polygenic risk scores to predict disease trajectories [15]. These successful efforts have primarily identified prognostic sub-phenotypes and clusters that carry an elevated cardiometabolic risk.

Nevertheless, the core challenge remains the unpredictable natural history of MASLD—a significant translational barrier that necessitates identifying the molecular determinants of severe outcomes and mortality. To address this knowledge gap, the present study was designed to define the molecular heterogeneity underlying MASLD clinical presentation, aiming to resolve different patterns of disease manifestation and identify sub-phenotypes that account for prognostic differences. We leveraged Latent Class Analysis (LCA) on a large UK Biobank (UKB) cohort to objectively assign individuals to distinct MASLD-phenotypic classes, followed by large-scale proteomic profiling. We focused on this molecular dimension because it directly reveals regulatory processes post-transcription. This approach unveiled distinct molecular subtypes with markedly different prognostic and survival outcomes. Using Cox proportional hazards (Cox PH models), we identified a validated subset of circulating proteins that facilitate the prediction of these prognostic subtypes, confirmed using an alternative proteomics platform in an external dataset. Critically, the differential mortality rates observed across these classes directly underscore that molecular heterogeneity impacts overall survival, providing a novel framework for risk stratification. This study is not an exercise in general population subtyping; rather, it uses the population as a canvas to definitively map the pathological boundaries and molecular drivers of mortality related to MASLD and associated cardiovascular complication.

## METHODS

### Rationale for population-wide LCA

To resolve the molecular heterogeneity inherent in MASLD, we employed a top-down LCA applied to the entire phenotyped cohort rather than a pre-filtered, disease-specific subgroup. This strategic design was predicated on the understanding that MASLD exists as a continuous spectrum of metabolic and inflammatory dysfunction rather than a binary clinical state. By eschewing rigid diagnostic thresholds at the discovery phase, we circumvented potential spectrum bias and captured sub-clinical metabolic shifts that often precede overt steatosis but significantly impact long-term mortality. Furthermore, this population-wide approach facilitated internal benchmarking by objectively identifying metabolically healthy reference classes within the same multidimensional space. This provided a mathematically rigorous baseline to quantify the degree of proteomic and prognostic deviation exhibited by high-risk MASLD endotypes. Ultimately, this unbiased framework enabled the discovery of hidden risk, revealing that individuals who do not yet meet traditional diagnostic criteria for MASLD can nonetheless harbour molecular endotypes associated with catastrophic mortality trajectories, thereby identifying a critical window for early clinical intervention.

### Study Design and Cohorts

The study comprised three phases utilizing data from four distinct cohorts.

Phase 1: MASLD phenotype and LCA. We characterized MASLD phenotype heterogeneity using LCA on 48,806 UKB participants (Cohort 1, *n* =48,806, 52% female, follow-up period 5718.5±307 days) with magnetic resonance imaging-based proton density fat fraction (MRI-PDFF) data. Classes were evaluated based on the updated MASLD definition traits (steatosis plus one of five cardiometabolic risk factors: arterial hypertension, T2DM /impaired glucose metabolism, obesity, hypertriglyceridemia, or decreased HDL-cholesterol), adjusted for alcohol consumption, age, and 10 principal genetic components (as a surrogate for ethnicity). All participants provided written, informed consent (NHS NRES Northwest reference: 16/NW/0274).

Cohort 1 was subsequently divided:

- Cohort 1a: participants (*n*=6,048) with both MRI-PDFF and blood proteomics data from the Pharma Proteomics Project (UKB-PPP), utilizing the Explore 3072 Proximity Extension Assay (PEA).
- Cohort 1b: participants with MRI-PDFF but no proteomics data (*n*=42,756).

Phase 2: Molecular Profiling and feature expansion. We examined the molecular heterogeneity of by profiling 2,923 proteins within each latent class. Differentially expressed proteins were assessed, and features were expanded via disease pathway analysis using protein enrichment.

Phase 3: Prognostic heterogeneity and signature identification. We identified subtypes with discordant survival rates using Cohorts 1a and 1b, and a distinct Cohort 2 (internal replication, *n*=44,252). Cohort 2 contained blood proteomics data but lacked MRI-PDFF for MASLD characterization.

External replication (Cohort 3, *n*= 22,913 participants from the Icelandic cancer project at deCODE) examined proteins previously linked to all-cause mortality, reusing publicly available summary data [15]. Samples were analysed using SomaScan v.4.0.

This research led to the identification of a proteo-survival signature for predictive score construction.

### Outcomes and Covariates

Complete details on the definition of outcomes of interest, covariates, exclusion criteria, and measurement of all-cause and cause-specific mortality are provided in the extended methods section (**Supplementary Material**).

### Protein measurements

Proteomic profiling of EDTA-plasma samples from 54,219 participants (baseline assessment) was performed with Olink Explore 3072 PEA (**Supplementary material** (extended methods).

Details on sample selection, processing, and quality control are described in earlier publications [16].

### Latent Class Analysis (LCA)

While traditional clustering methods rely on rigid spatial distances, LCA excels by acknowledging the inherent uncertainty and biological overlap within metabolic syndrome. By treating clinical phenotypes as hidden, unobserved entities rather than arbitrary groups, LCA effectively disentangles the complex interplay between obesity, hypertension, and fatty liver. This probabilistic framework moves beyond simple categorization, allowing researchers to uncover the subtle, underlying physiological mechanisms that define distinct patient subtypes, ultimately paving the way for a more nuanced and human-centred approach to precision medicine.

We employed generalized mixture models comprising two components: a measurement model defining the relationship between latent classes and observed indicators, and a structural model linking covariates and outcome variables to these classes. Latent class models were estimated using the following variables: Measurement variables (observed indicators): waist circumference, alcohol consumption, high-density lipoprotein (HDL) cholesterol, triglyceride levels, and three hepatic enzymes (ALT, AST, and GGT). Covariates: sex, age, and ethnicity. Dichotomous outcome variables: T2DM, hypertension, and SLD (steatotic liver disease). Details are provided in the extended methods section (**Supplementary Material**).

### Detailed Methodology

Full details on statistical analysis; latent class modelling; differential protein Expression (DEP) analysis; developing parsimonious protein scores (Lasso Regression); dynamic evaluation of predictive scores; ROC curve and survival analysis; pathway analysis; and causal mediation analysis are in the **Supplementary material** (extended methods).

## RESULTS

### Latent class analysis reveals sex-specific MASLD subtypes with distinct clinical profiles

The overall study design and key methodological phases are summarized in **Fig. 1**. We initiated the analysis by evaluating the phenotypic heterogeneity of Cohort 1, selected based on complete clinical and biochemical data for robust case analysis. Using LCA and phenotypes included in the new MASLD definition [2], we identified ten distinct latent clinical subgroups. Robust model fit, assessed by AIC (Akaike Information Criterion) and BIC (Bayesian Information Criterion), enabled partitioning of the spectrum into unique clinical scenarios. The relative strength of key phenotypic traits across the ten classes is detailed in **Fig. 2a**. These subgroups reflect underlying clinical heterogeneity and encompass diverse MASLD prevalences and multimorbidity profiles, each retaining a substantial sample size and follow-up period (females 5707±306/ males 5726±307 days) (**Fig. 2b**).

**Figure 1:**
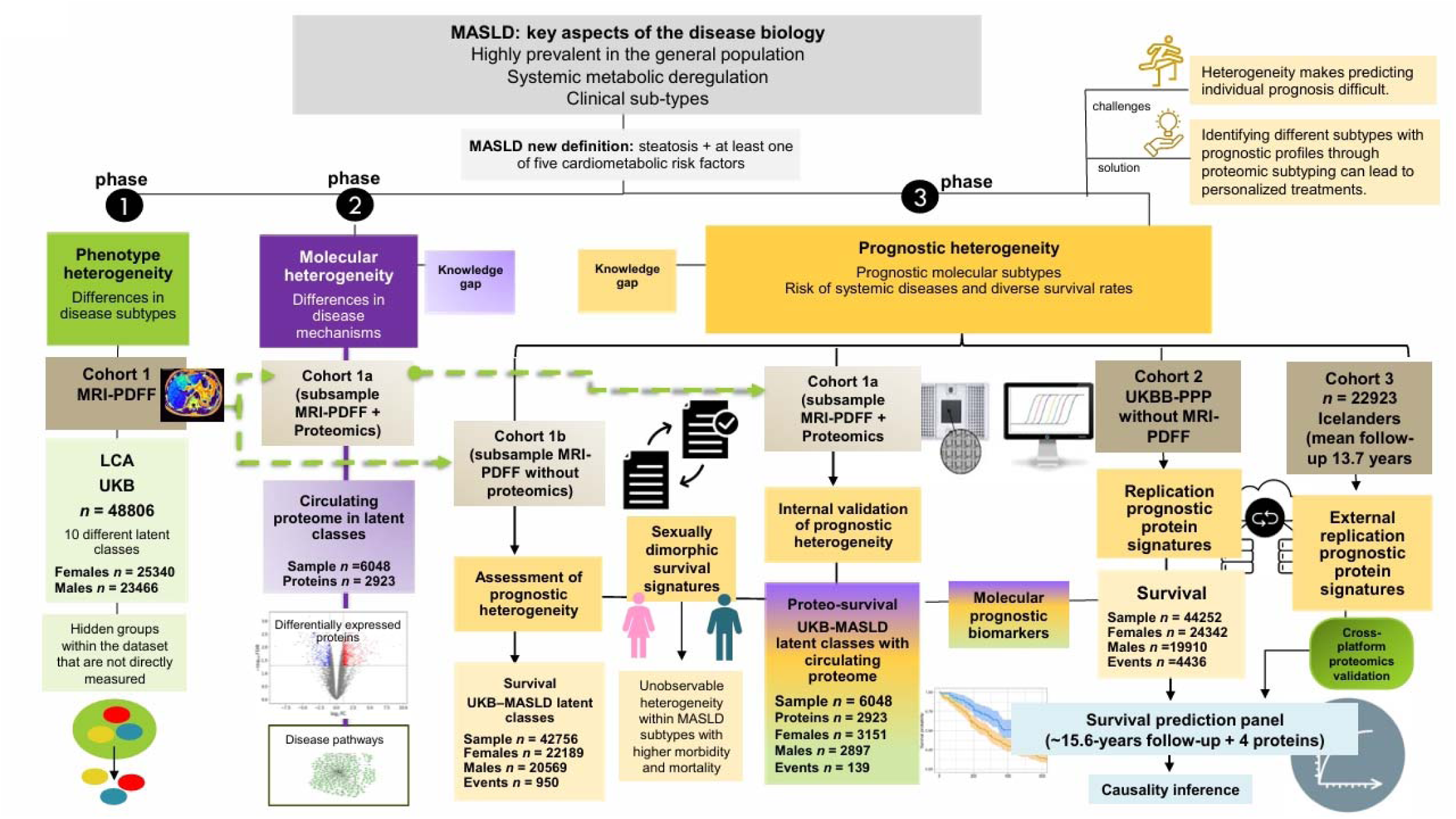
Schematic overview of study design, key methodologies, and phases. The study proceeded through three phases: **Phase 1 (Phenotype):** MASLD heterogeneity (*n*=48,806 UKB participants with MRI-PDFF characterized by Latent Class Analysis (LCA) of phenotypes, traits, and diseases. Cohort split: Participants with MRI-PDFF but without proteomics (*n*=42,756) and participants with MRI-PDFF and UKB-PPP proteomics (*n*=6,048). **Phase 2 (Molecular):** Molecular heterogeneity assessed by profiling 2,923 proteins within latent classes; differentially expressed proteins and disease pathways identified via enrichment analysis. **Phase 3 (Prognostic):** Prognostic heterogeneity explored by identifying MASLD subtypes with distinct survival rates (internal and external validation), leading to the proteo-survival signature and PS4 predictive score. PS4 components were implicated in the causal mediation of class effects on all-cause mortality.

**Figure 2:**
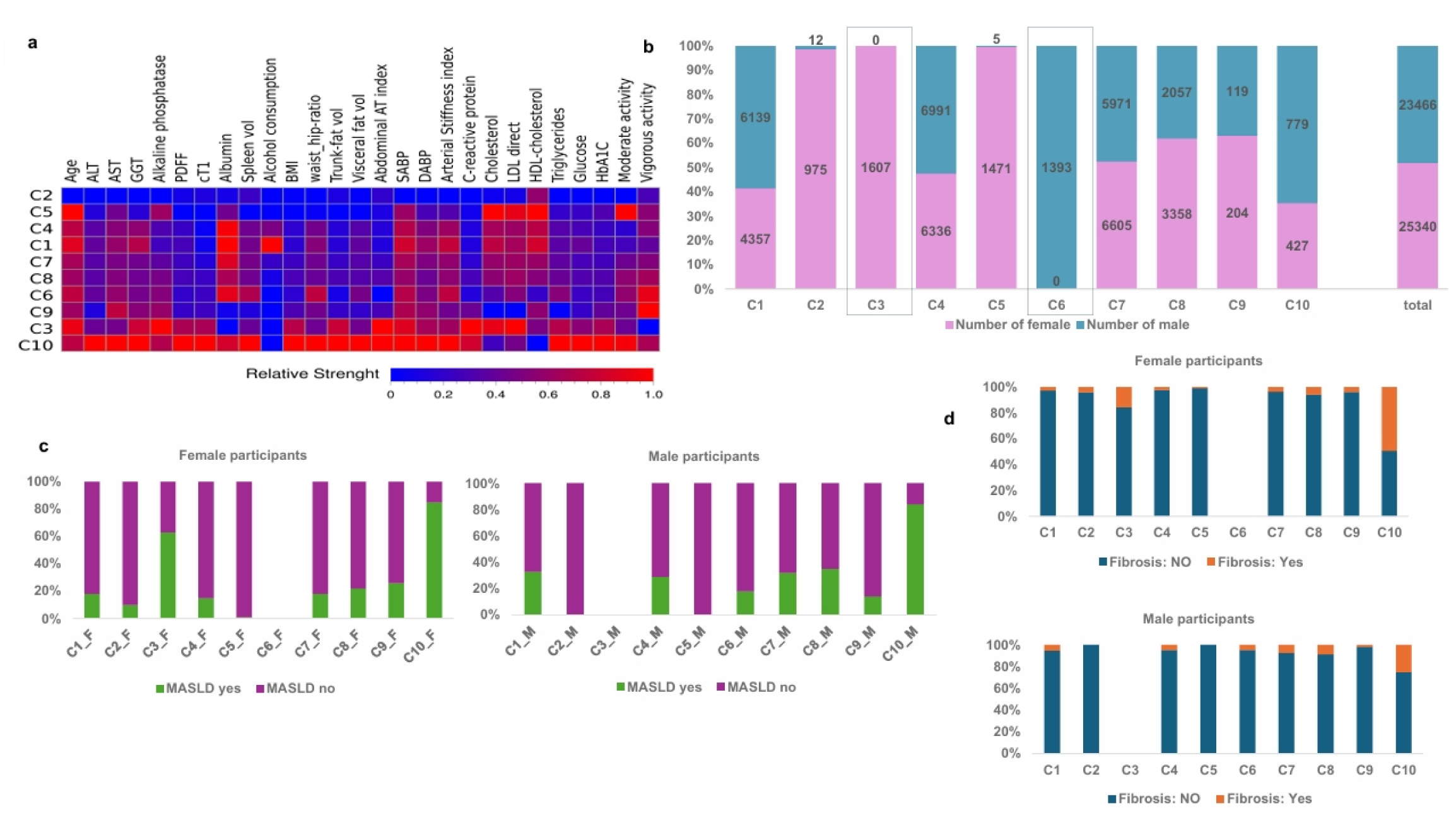
Latent class analysis reveals sex-specific MASLD subtypes with distinct clinical profiles. **a**. Relative strength of clinical and biochemical variables across LCA classes (means normalized 0 to 1 for comparison). **b**. Distribution of studied participants per class by biological sex. Black box highlights sex-exclusive classes. **c**. Prevalence of MASLD assessed by MRI-PDFF, stratified by sex within each class. **d**. Prevalence of liver fibrosis estimated using liver iron-corrected T1 (cT1), an MRI-based quantitative metric for hepatic fibro-inflammatory activity. Abbreviations: ALT/AST: alanine/aspartate aminotransferases; GGT: gamma glutamyl transpeptidase; PDFF: Proton density fat fraction; cT1: iron-corrected T1score (ms), indirect marker of hepatic fibro-inflammatory activity; BMI: body mass index; SABP/DABP: systolic/diastolic blood pressure; LDL direct: low-density lipoprotein cholesterol; HDL-cholesterol: high-density lipoprotein cholesterol; HbA1c: glycated haemoglobin; Physical activity defined as minutes/day of moderate (100–399) or vigorous (≥400 mins/day) activity based on mean acceleration values.

Striking sex-specific profiles were revealed by participant distribution (**Fig. 2b**): Class 3 comprised exclusively females, whereas Class 6 comprised solely males. MASLD prevalence, objectively assessed by MRI-PDFF, varied markedly across all classes (**Fig. 2c**). Class 10 exhibited the highest prevalence for both MASLD and liver fibrosis (**Fig. 2d**), consistently across both sexes. Specifically, MASLD prevalence in Class 10 was 85% (females) and 84% (males) (**Fig. 2c**); fibrosis (assessed by cT1) was 50% in females and 25% in males (**Fig. 2d**). Class 3 (female-only) represented the next high-risk subgroup (65% MASLD, 15% fibrosis). In stark contrast, the male-only Class 6 showed remarkably low prevalence (MASLD: 17%, fibrosis: 0.4%). Classes 2 and 5 (mostly female) demonstrated minimal or no steatosis, consistent with favourable metabolic health. Detailed quantitative class-specific features are in **Supplementary Table S1**. Despite including 10 principal genetic components as LCA covariates, residual ethnic heterogeneity was observed (**Supplementary Fig. S1**), particularly prominent in Class 9.

### Proteomic profiles uncover distinct molecular heterogeneity and associated pathways in Classes

Blood proteomics was performed on 6,048 baseline participants from the LCA subclasses with available proteomic data (females *n*=3,151; Cohort 1a) with available proteomic data from the UKB-PPP (**Supplementary Table S2**). This analysis used the Olink Explore 3072 PEA platform, measuring 2,923 unique proteins across eight thematic panels [16] to provide a broad molecular profile (**Supplementary Table S3**). Detailed differentially expressed proteins (DEPs) per class are in **Supplementary Table S4**, and the distribution of unique and shared proteins is illustrated in **Supplementary Fig. S2a** and **b**, respectively.

Figures 3a and 3c visually display protein abundance differences between the two high-risk MASLD subgroups (Class 3 and Class 10) and the remaining classes. These figures highlight DEPs in these two classes, which share the highest prevalence of MASLD with fibrosis. Enrichment analysis of biological processes (BP) revealed distinct underlying molecular mechanisms (Fig. 3b). Class 10 (both sexes) is primarily enriched for BP related to negative regulation of apoptosis and signal transduction, suggesting a strong cell-survival/proliferative phenotype. In contrast, the female-only Class 3 showed significant enrichment for chemotaxis and inflammatory response, highlighting immune-mediated disease activity. Furthermore, a substantial overlap exists: Class 10 and Class 3 share 94 upregulated and four downregulated proteins (Fig. 3d**-e**). To clarify shared mechanisms, we examined BP linked to common upregulated proteins across high-risk classes. Reactome pathway analysis revealed significant enrichment in key processes: CD163-mediated anti-inflammatory response, histidine catabolism, heme degradation, and interleukin-10 signalling (Fig. 3f; **Supplementary Table S5**). Functional interrogation of these shared upregulated proteins using the DisGeNET database showed an overrepresentation of severe clinical outcomes, including liver cancer, obesity, CVD, and immunomodulated disorders (Fig. 3g).

**Figure 3:**
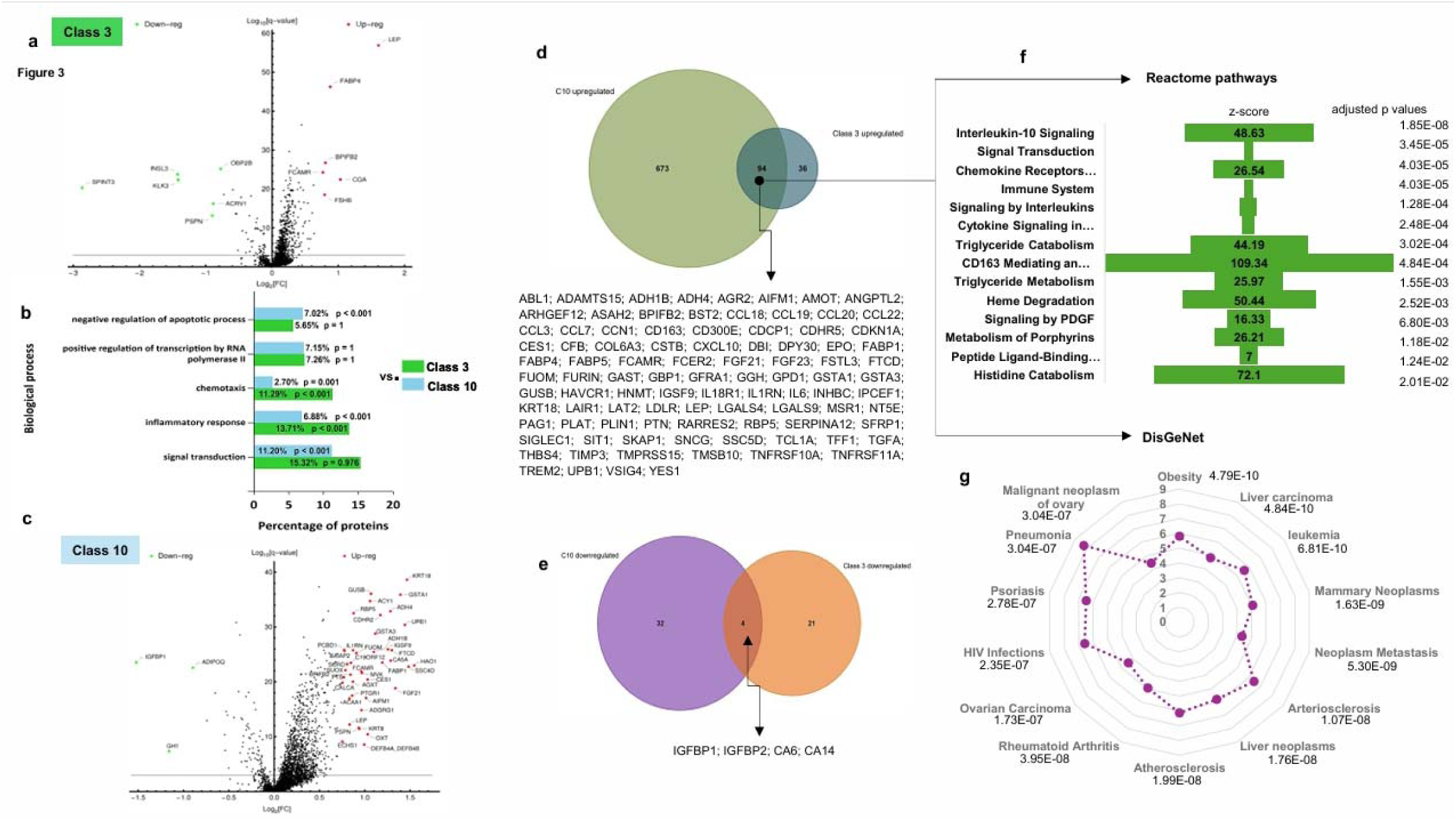
Proteomic signatures across MASLD clinical subgroups a, **c**. Volcano plots display proteins differentially expressed in Class 3 and Class 10 against all other classes (upregulated: log_2_(FC) ≥ 0.75, red; downregulated: log_2_(FC) ≤ −0.75, green), using a non-parametric Mann-Whitney U test and FDR q-value ≤0.001. **b**. Protein functional enrichment of Biological Processes (BP) in Classes 3 and 10 based on GO annotations, performed using FunRich (p-values adjusted by BH and Bonferroni). **d, e**. Shared differentially expressed (upregulated and downregulated) proteins in Classes 3 and 10. **f, g**. Reactome pathway analysis and DisGenet enrichment analysis for the set of shared proteins, performed using the Python library Enrich [33]. Enrichment results were considered significant with a BH-adjusted p-value threshold of <0·05.

### LCA defines MASLD prognostic subtypes with distinct survival outcomes

To investigate the prognostic implications of the MASLD subtypes, we structured the subsequent analysis into two distinct steps (Fig. 1). The initial exploratory step involved a survival analysis to assess prognostic heterogeneity among LCA classes. This analysis was performed in Cohort 1b (*n*=42,756; females *n*=22,189), which included MRI-PDFF data but lacked proteomic profiling. Over a mean follow-up of 5722.5±308 days (females: 5708±308, males: 5731±308), we captured 950 incident cases within the defined latent classes. Baseline characteristics for this group are summarized in **Supplementary Table S6**.

The analysis revealed a statistically significant association between all-cause mortality and sex (HR=1.48, 95%CI: 1.28-1.70, *p* = 9.51 × 10[[ and age (HR 1.12, 95%CI: 1.11-1.14, *p* = 1.06 × 10[¹[[). Subsequent Cox PH regression, adjusted for age (scaled to age 60), demonstrated that the high-risk subgroups—the female-specific Class 3 and Class 10—exhibited the highest all-cause mortality rates (Fig. 4a). These were compared against the minimal-risk reference group (Classes 1, 2, 4, 6), characterized by minimal MASLD prevalence. Figure 4b displays the age- and sex-adjusted distribution of Cox PH across all identified MASLD classes.

**Figure 4:**
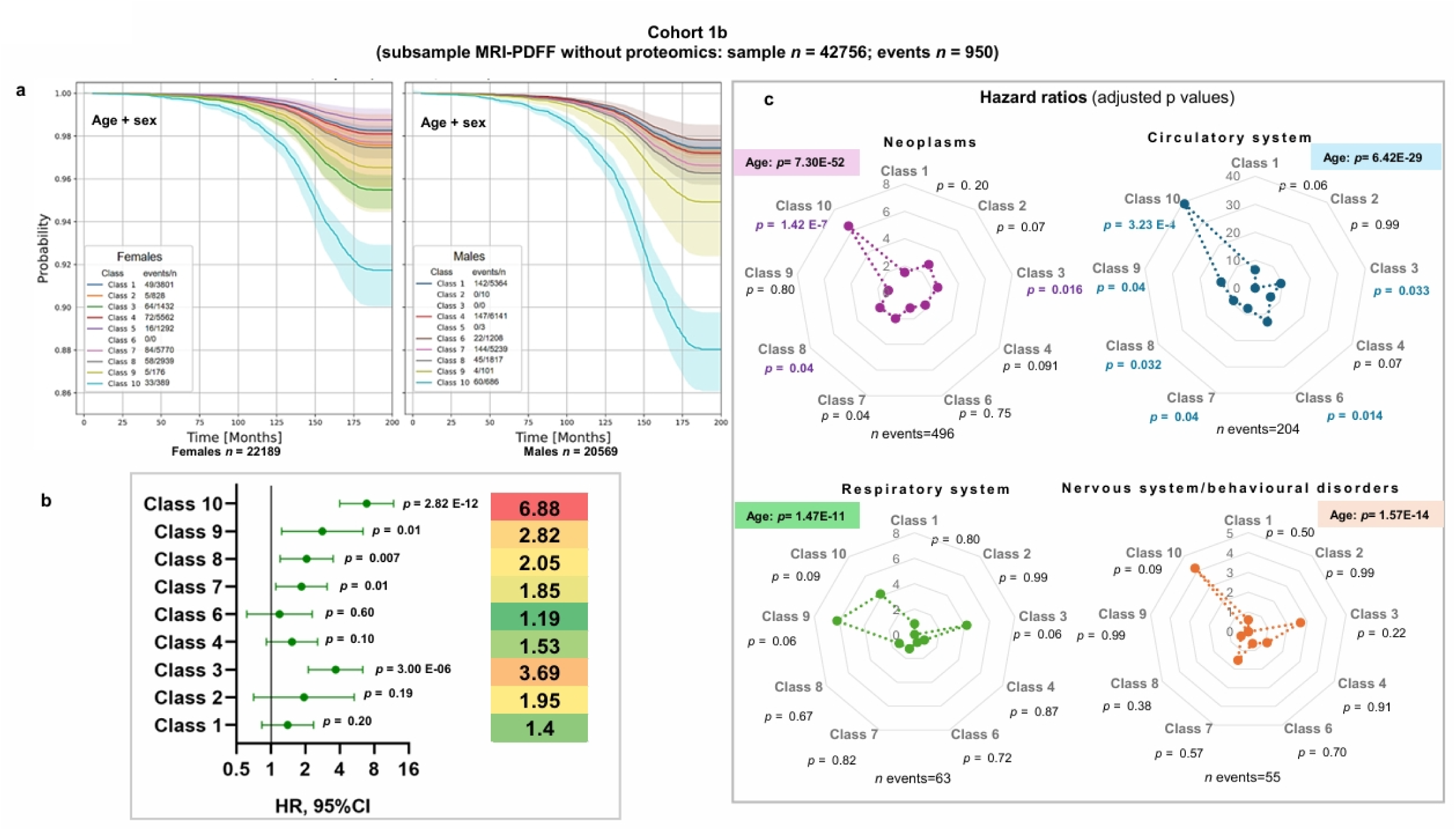
**LCA defines MASLD prognostic subtypes with distinct survival outcomes.** **a**. In cohort 1b, discriminatory capability of the all-cause mortality prediction model in females and males. The coloured shaded areas represent 95% confidence intervals. Cox PH model-based survival curves adjusted by age and ethnicity at 60 years old, stratified by sex: female (left panel), male (right panel), and classes. The survival curves illustrate the relationship between class membership and time to death in months (x-axis), with the y-axis representing the estimated probability of survival. The legend shows the number of events (deaths) and the number of observations (n) in each stratum. Classes without events are not shown. Class 5 establishes the baseline for comparative analysis, serving as the best-case scenario. The HR is used to calculate the relative risk of all other classes relative to the reference baseline. Cox PH regression revealed a statistically significant difference between sexes (males vs females, p = 9.51 × 10[[). Survival follow-up period (days): females 5709 ±308/ males 5732±308. **b.** In cohort 1b, discriminatory capability of the all-cause mortality prediction model applied to the entire sample (females and males) by classes, adjusting by age and sex. **c.** Age- and sex-adjusted HRs for major mortality causes (data field 40002), including neoplasms, and disorders of the circulatory, respiratory, and nervous systems.

Regarding mortality aetiology, the leading causes of death were neoplasms (52%), followed by diseases of the circulatory system (21.5%), respiratory system (6.6%), nervous system (5.8%), and digestive system (2.5%), with other internal causes accounting for the remaining 11.62%. Figure 4c displays the age- and sex-adjusted HRs for major mortality causes (data-field 40002). An increase in cause-specific mortality risk was most pronounced in Class 10. This class exhibited a significantly elevated risk for neoplasms (HR: 6.43, 95%CI: 3.21-12.86) and a markedly elevated, but less robustly estimated, risk for diseases of the circulatory system (HR: 39.28, 95%CI: 5.31 – 290.37) (Fig. 4c). Female participants of Class 3 also demonstrated a higher risk of cause-specific mortality due to neoplasms and cardiovascular disease, although the observed risk was lower than that of Class 10 (Fig. 4c). Comparative analysis of female-specific classes revealed distinct functional signatures (**Supplementary Fig. S3a, b**). Low-risk Class 5 (no MASLD, good survival) was significantly enriched for pathways related to glycoproteins, NODAL signalling, and hormone-ligand binding receptors, while prostanoid and sphingolipid metabolism were depleted. In contrast, proteins overexpressed in high-risk Class 3 were enriched in P53 transcriptional activation and cell cycle inhibitor P21. This was accompanied by depletion of DNA damage and replication checkpoints, pointing toward loss of genomic stability and molecular pathways linked to poor survival and high morbidity (e.g., malignancy).

### Development of a proteomic signature to predict survival risk across subclasses

To internally validate prognostic heterogeneity, we repeated the survival analysis in Cohort 1a (139 incident cases across the latent classes) defined by both MRI-PDFF and proteomic data. The mean follow-up was 5692 ±302 days (censored at death; females 5691±297/ males 5692±306). This reproduction confirmed the exploratory analysis: individuals in Classes 3 and 10 exhibited the highest all-cause mortality rates. We assessed model discriminatory capability using two Cox PH models (Fig. 5a, b). The initial model (Fig. 5a) was applied to all participants, stratified by sex and adjusted for age (normalized to age 60). The complementary model (Fig. 5b) was applied to the entire sample, adjusted for both age and sex.

**Figure 5:**
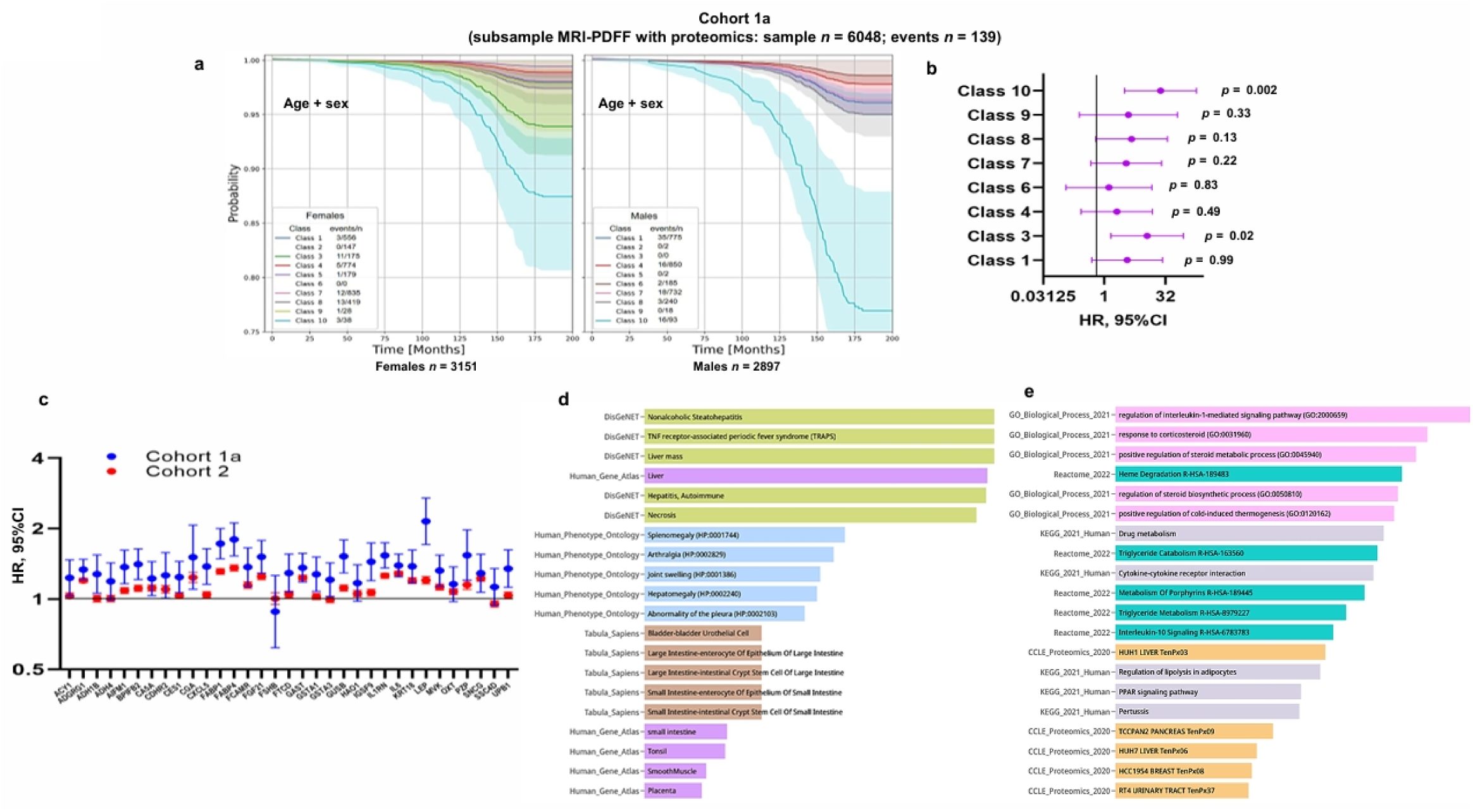
Development of a proteomic signature to predict survival risk across subclasses. **a-b.** Discriminatory capability of LCA subclasses in all-cause mortality prediction (Cohort 1a). Cox PH model was adjusted for age (normalized to 60) and ethnicity. **. Panel a:** Model stratified by sex (females/males). Follow-up: 5691±297 days (females) / 5692±306 days (males). **. Panel b:** Complementary model applied to the entire sample, adjusted for age and sex. **c**. Robustness of the 33-protein prognostic panel: Cox PH (HRs±95%CI) for all-cause death, adjusted for age and sex, across the entire sample (Cohort 1a and 2), following FDR correction. Proteins were normalized by z-scoring their NPX values. **d, e.** Cross-library analysis validation of the 24-protein signature used to build the final prediction model. The scale is also adjusted to fit the screen based on the maximum (complete data is in **Supplementary Figure 9**, including p value, q value, z-score, combined score, and number of proteins in the cluster).

After confirming the divergent prognostic profiles of Class 3 and Class 10, we explored their proteomic signatures to identify the most significantly deregulated proteins. From the 2,923 proteins assessed, we selected 33 differentially regulated proteins, 17 of which were common to both Class 3 and Class 10 (Fig. 5c). Using this list, survival analysis showed that 28 proteins retained a statistically significant positive association with all-cause mortality in the entire sample, even after FDR correction (Fig. 5c, blue dots; HRs 95% CI). The remaining five proteins (ADH4, FSHB, HAO1, OXT, and SSC4D) lacked a statistically significant association with mortality.

### Validation of Prognostic Protein Markers

To validate the robustness and generalizability of the prognostic markers, we conducted a replication analysis in an expanded, independent UKB cohort (Cohort 2, *n*=44,252; females *n*=24,342). This cohort included proteomic data but lacked MRI-PDFF confirmation of MASLD. The replication group, with a mean follow-up of 5418±971 days (females: 5490±851/ males: 5330±1093), utilized the 33-protein signature to assess risk. This approach revealed that 24 of the 28 previously prognostic proteins exhibited statistically significant HRs consistently in the same direction as the discovery cohort, though attenuated (Fig. 5c, red dots). Based on these robust results, this 24-set was selected to build the protein prediction score (**Supplementary Table S8**). Cross-library analysis of the 24-protein signature revealed enrichment (Fig. 5d, e; **Supplementary Table S9**) for key biological processes, enriched clinical phenotypes and tissue enrichment: liver is the most overrepresented tissue for the signature (Fig. 5d).

### Protein score demonstrates sustained predictive performance for all-cause mortality

To develop an optimal prognostic tool that avoids colinearity among the 24 proteins, we employed LASSO regression within Cohort 1a to determine a parsimonious score for all-cause mortality prediction (up to 10 years). Based on BIC best-fit, the resulting score (PS4) incorporated four key proteins: FTCD, FABP1, ADGRG1, and GAST (Fig. 6a). As Cohort 2 lacks MRI-PDFF, the non-invasive FIB-4 index (a predictor of major adverse liver and all-cause mortality outcomes [3,17]) was used as a baseline comparator. FIB-4 (≥2.67) has also been associated with the prediction of all-cause mortality in various populations [18–21], including patients with heart disease irrespective of MASLD status [18]. The predictive performance of two models (PS4+FIB4+sex+age and PS4+sex+age) was compared against a baseline model (FIB4+sex+age) using the area under the ROC curve (AUC). Throughout all time points analysed, PS4+sex+age consistently outperformed FIB4+sex+age, with no significant improvement observed from including FIB-4 (**Fig.6a**). **Fig.6b-c** presents ROC curves for mortality at 1500 days (AUC 0.751±0.018) and 2000 days (AUC 0.757±0.016), indicating that PS4+sex+age significantly outperforms FIB4+sex+age in predicting the outcome with a greater AUC, exceeding 0.050 difference. Analysis of protein and gene expression public datasets identifies the liver as the main source of circulating proteins associated with all-cause mortality (Fig. 6d**–e**). **Supplementary Fig. S4** presents ROC curves for each protein of the PS4 at the same time points. GAST and ADGRG1 showed marginally but not significantly better performance, particularly at 1,500 days.

**Figure 6:**
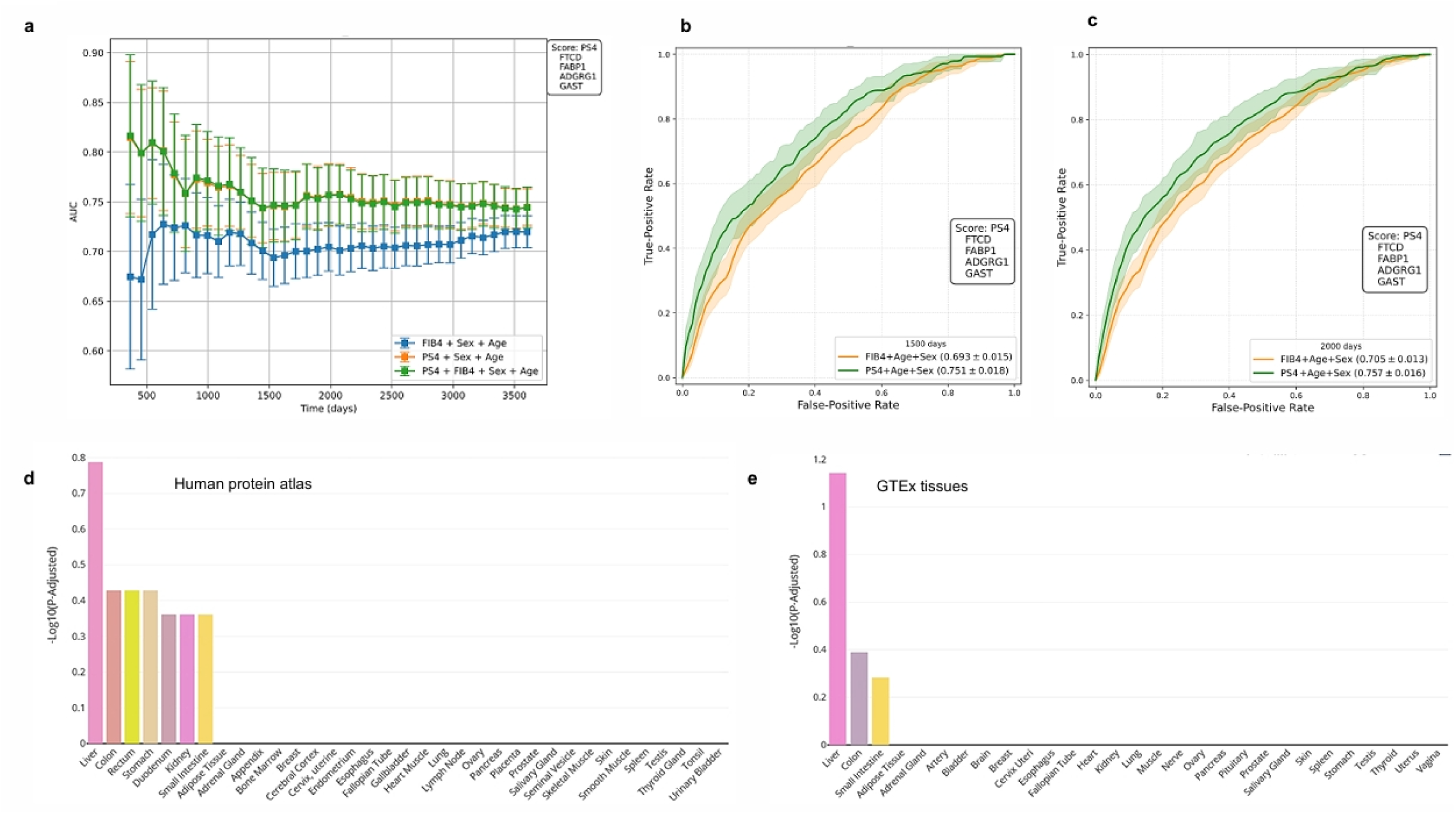
Protein score demonstrates sustained predictive performance for all-cause mortality. **a**: AUCs for all individuals of cohort 2, *n* = 44,252. Results are shown as AUC±95%CI at each time point of follow-up in days. Survival follow-up period (days), females: 5490±851/ males 5330±1093. **b, c**: ROC curves for all-cause deaths within 1500 and 2000 days for models described in the legends are shown in panels b and c, respectively. PS4 stands for the protein score obtained by LASSO with BIC, using the 24 proteins that were replicated across the cohorts, as described in the main text. **d, e**. Enrichment analysis of tissue expression of the selected proteins (FTCD, FABP1, ADGRG1, GAST) using the tissue enrich tool available at https://tissueenrich.gdcb.iastate.edu/. Histogram plot options: p-values adjusted.

### Replication and validation of the protein signature in the Icelandic cohort

To assess the generalizability and clinical utility of the prediction model, we performed a second replication study. This involved testing the selected proteins in an external population sample from the deCODE dataset (including the Icelandic cancer project) [15] (**Supplementary Table S10**). The deCODE cohort (cohort 3, *n*=22,923 participants) provided essential survival and cause-of-death data. Importantly, the proteomics data were generated using a different high-throughput platform, ensuring a robust, independent evaluation of the findings. Of our 24 proteins, 18 matched those tested by Eiriksdottir et al. [15], with LEP, KRT18, AIFM1, and FCAMR showing no significant impact on all-cause mortality (all p > 0.16). After excluding these four, LASSO analysis of the remaining 20 proteins yielded similar results and identified the same PS4 protein set (FTCD, FABP1, ADGRG1, GAST). Model performance remained comparable, as shown by similar AUCs in **Supplementary Fig. S5.**

### Mediation analysis of proteomic signatures on mortality

We used Cox PH mediation analysis with bootstrapping to determine if the relationship between Classes 3 and 10 and all-cause mortality was mediated by the four PS4 proteins (FTCD, FABP1, ADGRG1, and GAST), one at a time. Both class exposures remained significantly linked to higher mortality, even after adjusting for age and sex, with PS4 remaining an independent predictor. Decomposition revealed a partial mediation role for PS4: around 20% of the exposure–mortality association was mediated (significant indirect effect by bootstrap), though the substantial direct effect remained significant (**Supplementary Table S11**). In specific class-protein analyses, FABP1 mediated 20.7% of the effect in Class 3, while ADGRG1 mediated 20.1% in Class 10. The PS4 aggregate showed similar indirect effects across the classes, mediating 14.6% in Class 3 and 23.8% in Class 10 of the overall association (Fig. 7). Mediating roles in intermediate phenotypes are fully described in the Supplementary Material (Supplementary Table S12, Supplementary Fig S6).

**Figure 7:**
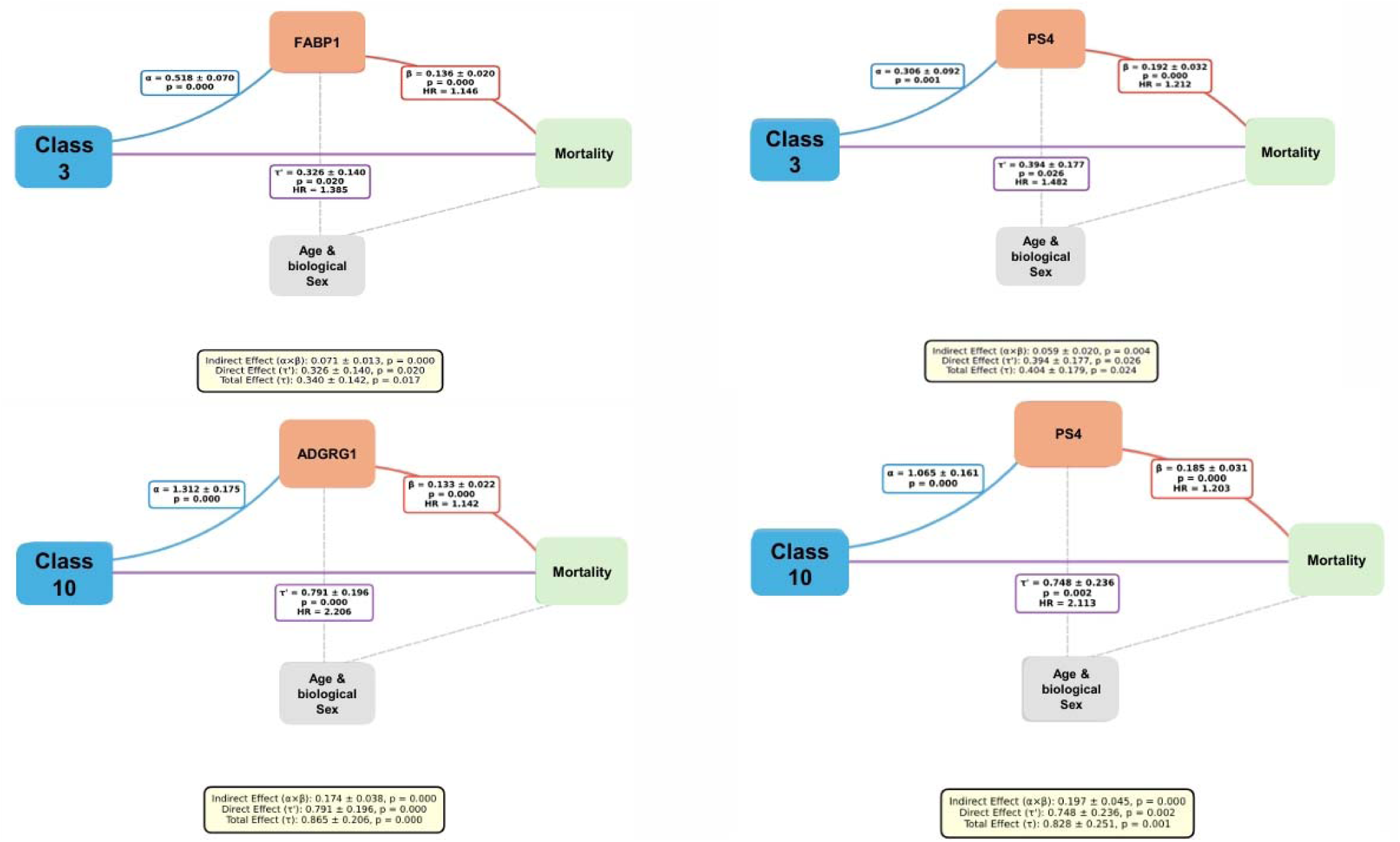
**Mediation analysis and causality assessment** Cox PH mediation analysis with bootstrapping to determine whether the relationships between classes 3 and 10 and all-cause mortality were mediated by PS4 and the selected proteins: FABP1 and ADGRG1. We adjusted all models for age and sex a priori. All analyses were implemented in Python (StatsModels) with custom utilities for mediation and diagnostics.

## DISCUSSION

Our successful stratification of MASLD subtypes into distinct clinical scenarios underscores the complex and multifaceted nature of the disease. This complexity is highlighted by the high prevalence (>85%) among individuals presenting with the full spectrum of cardiometabolic risk factors [6]. Crucially, we observed a pronounced sexual dimorphism in disease severity: advanced fibrosis was present in 50% of females versus only 20% of males. Furthermore, LCA reinforced this sex-specific burden by identifying a separate female-only class where liver fibrosis reached 15%. These findings collectively emphasize the substantial clinical heterogeneity of MASLD, necessitating stratification that extends beyond traditional risk factors to incorporate biological sex directly.

To define the underlying molecular signatures driving this clinical heterogeneity, we performed in-depth blood proteomic profiling. Although the high-risk MASLD classes (Class 3 and Class 10) shared several overrepresented proteins, their molecular signatures significantly diverged. Notably, proteins common to both high-risk classes were significantly enriched for tissue repair and scavenging mechanisms, exemplified by the CD163 scavenger receptor (highly expressed on M2 macrophages) [23].

The identified heterogeneous molecular signatures were directly associated with clinical outcomes, demonstrating clearly divergent survival patterns over the >15-year follow-up. After accounting for age and sex, Classes 3 and 10 exhibited the highest all-cause mortality rates (HR: 3.69 and 6.88, respectively). Critically, these mortality risks were predominantly driven by neoplasms and CVD in both high-risk classes. In the primary care setting, where MASLD is most frequently managed, a patient’s primary risk is not limited to end-stage liver disease. The systemic liver-related risk in MASLD encompasses accelerated atherosclerosis and oncogenesis, fuelled by hepatic metabolic dysfunction. By capturing all-cause mortality, our PS4 score reflects the comprehensive biological burden of the disease, providing a more clinically representative assessment of patient risk than liver-specific mortality alone, which represents only the terminal fraction of the disease spectrum. Furthermore, focusing exclusively on liver-related death in a population-wide cohort would lead to a significant underestimation of the disease’s actual impact on public health, as the metabolic spillover from a dysfunctional liver often reaches a fatal threshold within cardiometabolic or oncological systems well before the onset of cirrhosis-related failure.

Building on this prognostic divergence, we leveraged proteomic signatures to develop a robust, clinically translatable model for predicting survival risk and exploring biological causality. LASSO regression yielded an optimal four-protein signature (PS4) with strong prognostic discrimination. This was robustly validated in a separate, larger replication cohort with a higher number of mortality events, showing consistent effects and directionality. Furthermore, PS4 selection was replicated in an external cohort using a different ethnic background and proteomic platform [16], thus establishing its generalizability.

The four proteins comprising the PS4 signature (FTCD, ADGRG1, FABP1, and GAST) illustrate the key functional pathways linking MASLD to systemic risk. FTCD (folate pathway) [24] and FABP1 (lipid transport) connect hepatic metabolic stress and lipid overflow directly to cellular damage [25]. ADGRG1 [26,27] and GAST (Gastrin) [28] are associated with pathways regulating immune cell trafficking, vascular integrity, and proliferation. Collectively, the signature components directly contribute to the systemic development of CVD and neoplasm risk, the primary mortality drivers in our high-risk classes.

When benchmarked against established non-invasive scores (e.g., FIB-4), the PS4 signature significantly outperformed them, achieving a consistently high AUROC of 0.75 for mortality prediction at both 1,500 and 2,000 days. Analysis confirmed the PS4 proteins are predominantly released from the liver, highlighting the direct role of MASLD/MASH pathogenesis in driving mortality risk. While PS4 prognostic performance is competitive with tools like liver elastography in high-risk settings [29–31], its fundamental strength lies in its biological underpinning and causal relevance. Unlike liver stiffness (a structural measure), our signature identifies proteins that are active effectors within disease pathways. This evidence suggests PS4 captures independent biological information related to active progression, thereby informing prognosis and pointing toward actionable therapeutic targets. To dissect the functional role of the PS4 signature in driving mortality, we performed formal mediation analysis. This analysis demonstrated that the PS4 proteins significantly mediate the association between the high-risk classes and all-cause mortality, accounting for 10 to ∼24% of the total mortality effect. Importantly, FABP1 mediated 20.7% of the effect in the female only class 3, while ADGRG1 mediated 20.1% in both sexes class 10.

When analysing the PS4 signature’s mediation across individual cardiometabolic traits, we found its component proteins capture a pathway of systemic risk. For instance, FTCD was a prominent mediator across multiple endpoints (including fibrosis, steatotic liver disease, arterial hypertension, and T2DM), with ADGRG1 also contributing meaningfully to these outcomes. These results suggest that the biological processes reflected by the PS4 signature are not merely downstream consequences of established pathology, but rather novel, active mechanisms of systemic deterioration originating in the diseased liver.

Collectively, the identification of prognostically divergent MASLD subtypes, coupled with a highly validated, non-invasive, proteomic signature for all-cause mortality, fundamentally moves beyond simple steatosis grading to establish a clear and quantifiable link between hepatic disease severity and systemic, long-term patient outcome.

This strategy aligns with the growing body of literature utilizing proteomic signatures to refine MASLD prognosis. Specifically, recent biomarker scores [32–36] have successfully predicted advanced fibrosis, while others have linked individual proteins directly to long-term systemic complications [37,38], or predicted drug response [39]. This distinction—moving from predicting an intermediate marker (fibrosis) to predicting the definitive outcome (mortality), explaining some causal mediator effect—is what makes the PS4 score a major advance.

A primary limitation is the potential selection bias within the UKB: the cohort population is typically healthier and less diverse than conventional clinical populations. This is significantly counterbalanced by the successful replication of the MASLD prognostic signature in the independent Icelandic cohort, underscoring its robustness and generalizability. Because MASLD management often occurs in primary care, assessing these signatures in the general population is clinically relevant; however, caution is warranted when extrapolating globally. Additionally, the PS4 proteins were selected by unsupervised LASSO which, probably due to collinearity, excluded candidates worthy of future testing and previously associated with MASLD.

In conclusion, the implementation of a top-down LCA proved instrumental in partitioning the continuous cardiometabolic–MASLD spectrum into biologically distinct endotypes, thereby significantly enhancing the precision of risk stratification. This strategy enabled the discovery of natural, data-driven groupings that are unconstrained by the limitations of conventional diagnostic labels. By interrogating the entire UKBB cohort, our model successfully delineated the threshold where physiological normalcy transitions into pathology. Consequently, MASLD is no longer viewed as a static, isolated organ disease, but as a dynamic set of molecular clusters that have diverged from a metabolically healthy baseline. Crucially, the primary intent of this work is not to introduce a novel non-invasive test intended to supersede established fibrosis scores in routine clinical practice. Rather, our findings establish a robust mechanistic framework that characterizes the molecular events defining the MASLD state and the pathways through which they dictate systemic outcomes. By using all-cause mortality as an endpoint in a population-wide cohort, we have mapped the pathological boundaries and molecular drivers of MASLD-related survival. In this context, the PS4 proteins function as biological reporters of active disease processes. While their predictive performance exceeds that of existing models, their fundamental value lies in identifying the 10–24% causal fraction of mortality attributable to specific metabolic and immune-vascular pathways activated in high-risk MASLD subclasses. This distinction separates the PS4 from markers of structural liver scarring, positioning it as a fundamental tool for deciphering disease biology and identifying mechanism-specific therapeutic targets.

## Data Availability

All datasets generated and analysed during the current study are available in the Supplementary Material accompanying this manuscript (https://doi.org/10.5281/zenodo.17764543). The code used for the analytical procedures (Latent Class Analysis, proteomic score derivation, and survival modelling) is not original and is referenced within the Methods section.

## Acknowledgements

This research has been conducted using the UK Biobank Resource under Application Number 485675

## Authors’ contributions

**SS and CJP** designed the study. LD, SS and CJP analysed the results. LD, SS and CJP drafted the manuscript. LD and CJP contributed to the acquisition and analysis of data. CJP provided statistical support. LD, SS and CJP revised and approved the final version of the manuscript. SS and CJP are the guarantor of this work, with full access to all data and analyses related to the content of this article.

## Supplementary Material

Due to the **substantial volume** of the data contained within the **supplementary tables** (Supplementary Tables 1-12), these files have been deposited in the **Zenodo** public repository to ensure permanent access and ease of download. The complete dataset can be accessed using the identifier: [https://doi.org/10.5281/zenodo.17764543].

## Extended methods

### Definition of outcomes of interest and covariates

The primary outcome was MASLD. MASLD was defined using magnetic resonance imaging-derived proton density fat fraction (MRI-PDFF) as a non-invasive, quantitative measure of steatosis (data fields 21088 and 24352); liver fibrosis was assessed using liver iron corrected T1 (cT1) (data field 40062). We used the International Classification of Diseases, 10th Revision (ICD-10) system to define the other outcomes of interest.

The studied variables include demographic characteristics such as biological sex (variable 21), age at recruitment (data field 21022) and age at death (data field 40007), anthropometric features such as weight (data field 23098), body mass index (BMI) (data field 23104), visceral adipose tissue volume (VAT) (data field 22407), visceral fat volume (data field 21085), adjusted T/S ratio (data field 22191), total trunk fat volume (data field 22410), weight-to-muscle Ratio (data field 22433) and waist circumference (data field 48) to assess general and central body adiposity. Biochemical tests commonly used in the clinical setting to stratify risk for MASLD including aminotransferase (alanine, ALT [data field 30620] and aspartate transaminase, AST [data field 30650]), gamma-glutamyl transferase (γGT) (data field 30730) and alkaline phosphatase levels (AP) (data field 30610), as well as total bilirubin (data field 30840) and liver (data field 21080) and spleen volume (data field 21083) were assessed to evaluate liver involvement. The assessment of fasting glucose (data field 30740) and glycated haemoglobin (HbA1c) (data field 30750) levels were undertaken to characterise impaired glucose metabolism. The evaluation of total cholesterol (data field 30690), high-density lipoprotein cholesterol (HDL-C) (data field 30760), low (data field 23405) and very low-density lipoprotein (data field 23403) cholesterol (LDL-C and vLDL-C), triglycerides (data field 30870), apolipoprotein A (data field 30630) and B (data field 23439) was conducted to assess lipid metabolism. Furthermore, serum creatinine (data field 30700) and urea (data field 30670) levels were evaluated as surrogates of kidney function. C-reactive protein (data field 30710) was utilised as an indicator of systemic inflammation. Systolic blood pressure (data field 4080) and diastolic (data field 4079) blood pressure, arterial stiffness (data field 21021) was utilised as indicators of CVD. Physical activity: this category contains data from the touchscreen questionnaire on the reported type and duration of physical activity (including walking, DIY, moderate and vigorous physical activity, strenuous sports, etc); duration of vigorous activity (data field 914) and duration of walks (data field 874) expressed as minutes/day. Amount of alcohol drunk on a typical drinking day (data field 20403) and alcohol drinker status (data field 20117).

### Ethnic background

Information on ethnicity is under the data field 21000. Distribution is as follows: white *n* = 555149; mixed *n*= 3423; Asian or Asian British *n*= 10795; Black or Black British *n*= 8670; Chinese *n*=1803; other ethnic group *n*= 4979; do not know *n*= 236; prefer not to answer *n*= 1874.

### Measurement of all-cause and cause-specific mortality

Death notifications on a regular basis through linkage to national death registries (data field 40023); the main cause of death for each participant is identified by the ICD10 code. Survival analysis included age at death (data field 40007) and sex. Participants from the UKB were followed for all-cause and cause-specific mortality until 31 May 2024 (England & Wales, data provider: NHS England) and 31 December 2023 (Scotland, data provider: NHS Central Register, National Records of Scotland). The data UK Biobank receives from the death registry includes the date of death and the primary and contributory causes of death, coded using the ICD-10 system (data field 40001; description: underlying (primary) cause of death). The death data available in a main dataset consists of the data-fields in Category 100093, that contains the following information: age at death (data-field 40007) - a derived data-field calculated as the interval between date of birth and the date of death, and description of cause of death (data-field 40010) - free-text cause of death information from the death certificate coded according to the ICD-10 (Chapter I Certain infectious and parasitic diseases; Chapter II Neoplasms; Chapter III Diseases of the blood and blood-forming organs and certain disorders involving the immune mechanism; Chapter IV Endocrine, nutritional and metabolic diseases; Chapter V Mental and behavioural disorders; Chapter VI Diseases of the nervous system; Chapter VII Diseases of the eye and adnexa; Chapter VIII Diseases of the ear and mastoid process; Chapter IX Diseases of the circulatory system; Chapter X Diseases of the respiratory system; Chapter XI Diseases of the digestive system; Chapter XII Diseases of the skin and subcutaneous tissue; Chapter XIII Diseases of the musculoskeletal system and connective tissue; Chapter XIV Diseases of the genitourinary system; Chapter XV Pregnancy, childbirth and the puerperium; Chapter XVII Congenital malformations, deformations and chromosomal abnormalities; Chapter XVIII Symptoms, signs and abnormal clinical and laboratory findings, not elsewhere classified; Chapter XX External causes of morbidity and mortality; Chapter XXII Codes for special purposes).

### Protein measurements

Proteomic profiling of EDTA-plasma samples from 54,219 UKB participants from the baseline assessment was performed with Olink Explore 3072 PEA, measuring 2,941 analytes representing 2,923 unique proteins. The strategy for the selection of samples for proteomic profiling was designated by members of the UKB-PPP consortium. The average intra- and inter-plate %CVs are very low, and assay detectability was over 89% across panels. Over 92% of the samples have passed QC for all assays. We used the reported normalized protein expression (NPX) values. Calculation of Normalized Protein eXpression (NPX) values was performed stepwise as described in data field 30900 using a two-step normalization approach consisting of within-batch and across-batches intensity normalization. Briefly, the log2 Ratio was calculated of counts of each assay of each sample to the counts of the extension control, and the assay-specific median value of the plate controls was subtracted. This provided plate control (PC) normalized NPX values. Next, the assay-specific plate median NPX value was subtracted, and the batch-specific median NPX value for each assay was added to account for effects within each batch.

### Latent class analysis

We employed generalized mixture models, which comprise two components: a measurement model that defines the relationship between the latent classes and their observed indicators, and a structural model that links covariates and outcome variables to these latent classes.

For the estimation of the latent class models, we utilized the following variables:

- Measurement variables (observed indicators): waist circumference, alcohol consumption, high-density lipoprotein (HDL) cholesterol, triglyceride levels, and three hepatic enzymes (ALT, AST, and GGT).
- Covariates: sex, age, and ethnicity.
- Dichotomous outcome variables: T2DM, hypertension, and SLD (steatotic liver disease).

We utilized several variables directly from the UKBB, including waist circumference, HDL cholesterol, triglyceride levels, ALT, AST, GGT, sex, and age. To maximize the number of subjects and improve subsequent analyses, certain outcome variables were redefined using a broader set of criteria and related data fields from the database.

Specific outcome variable definitions: the following criteria were used to define the outcome variables, including individuals with a formal diagnosis and those with clinical parameters compatible with the condition: T2DM: Individuals were classified as having T2DM if they met any of the following criteria: Diagnosis corresponding to ICD-10 categories E11, E13, or E14 (Data Field 41270). A positive self-reported diagnosis for the disease (Data Field 130709). Clinical parameters compatible with diabetes, defined as: Glucose (Data Field 30740) ≥5.6 mm/L; HbA1c (Data Field 30750) ≥39mm/L. Similarly, individuals were classified as having hypertension if they met either of the following criteria: diagnosis corresponding to the ICD-10 categories I10, I11, I12, or I13 (Data Field 41270). A positive self-reported diagnosis for hypertension (Data Field 131287). We also included individuals with clinical parameters consistent with hypertension, even in the absence of a formal diagnosis. In this regard, the following criteria were used: Systolic Arterial Blood Pressure (SABP) (Data Field 4080) ≥130 mmHg; Diastolic Arterial Blood Pressure (DABP) (Data Field 4079) ≥85 mmHg.

For SLD, we utilized a definition incorporating both formal diagnosis and clinical imaging parameters. Formal Diagnosis: Individuals were classified as having SLD if they had a diagnosis corresponding to the ICD-10 category K76.0 (Data Field 41270). Imaging Parameters (Subclinical Disease): To recruit subjects with subclinical disease, we incorporated two liver Proton Density Fat Fraction (PDFF) measurements from UKBB at Instance 2: Average PDFF: The average PDFF was derived from at least three (and up to nine) regions of interest in the liver (Data Field 24352). Liver Fat Percentage: The measure of total liver fat percentage (Data Field 40061).

The two available liver PDFF measurements captured different cohorts: the first measure (Data Field 24352) assessed 46,854 participants, while the second measure (Data Field 40061) assessed 40,483 participants. Given the substantial overlap between the two measurement groups, we implemented a simple linear imputation strategy to maximize our study cohort size.

For participants assessed by both methods, we constructed a linear model to define the relationship between the two PDFF measures. This model was used to extend the cohort as follows: Imputation: If the variable was missing, we imputed its value using a prediction generated by the linear model, provided that had a valid measurement. Exclusion: If both were missing for a specific subject, the PDFF output remained missing, and that subject was subsequently excluded from this analysis cohort.

We addressed missing and categorical data for alcohol consumption as follows. We excluded subjects who chose not to report on their alcohol consumption status (Data Field 1558, coded as value −3). Scoring and Mapping: To create a quantitative measure of alcohol consumption frequency, we established a proportional score based on the categorical responses in Data Field 1558 using the following mapping= {1->8, 2->4, 3->2, 4->1, 5->0.5, 6->0}.

For ethnicity, we controlled for the first ten principal components (PCs) derived from Data Field 22009.

Fibrosis and FIB-4: other variables were defined and calculated within the cohort: Fibrosis Status (fibrosis): A dichotomous variable defined based on the continuous cT1 measure. Fibrosis yes: if cT1 >800/ fibrosis no cT1 <800. FIB-4 Index (FIB4): a clinical index calculated using standard formula inputs: Platelet Count (Data-Field 30080); age (Data-Field 21022); AST (Aspartate Aminotransferase) (Data-Field 30650); waist-to-Hip Ratio (waist\hip-ratio).

### Exclusion Criteria

For all subsequent analyses, we applied stringent exclusion criteria to ensure the study cohort was free from conditions that could confound the proteomic signatures, particularly those related to severe infection or advanced liver disease.

### Exclusion by Serostatus and Specific Diagnosis

We excluded participants who were seropositive for the following conditions: Hepatitis B Virus (HBV) (Data Field 23060); Hepatitis C Virus (HCV) (Data Field 23061); Human Immunodeficiency Virus (HIV) (Data Field 23064). Additionally, we excluded participants with a diagnosis in the ICD-10 range of B15.9 to B25.9 (Data Field 41270), which covers a range of viral diseases including acute hepatitis and cytomegalovirus.

Exclusion by Disease/Condition (ICD-10 Categories)

We also excluded individuals with one or more diagnoses of the following severe diseases or conditions (based on ICD-10 categories in Data Field 41270):

- Advanced liver pathology:
- `o` C22.0: Liver cell carcinoma
- `o` K70: Alcoholic liver disease (all sub-codes)
- `o` K71: Toxic liver disease (all included sub-codes)
- `o` K72: Hepatic failure, not elsewhere classified (all sub-codes)
- `o` K73: Chronic hepatitis, not elsewhere classified (all sub-codes)
- `o` K74: Fibrosis and cirrhosis of liver (all sub-codes)
- `o` K75: Other inflammatory liver diseases (all sub-codes)
- `o` K76: Other diseases of liver (excluding K76.0, as used for SLD definition)
- `o` K77: Liver disorders in diseases classified elsewhere.
- Related conditions:
- `o` E83: Disorders of mineral metabolism (E83.0, E83.1)
- `o` F10: Mental and behavioural disorders due to use of alcohol (F10.0-F10.4)
- Transplant status:
- `o` Z94.4: Liver transplant status

### Latent class modelling

Applying all criteria, we established a final analysis cohort of 48,806 individuals from the general population with no missing values in the measurement, covariate, or outcome variables.

This complete data matrix was analysed using StepMix, a Python package designed for the pseudo-likelihood estimation of generalized mixture models. We employed the one-step approach, utilizing the expectation-maximization (EM) algorithm to simultaneously estimate parameters for both the measurement and structural models.

Some measurement variables are continuous but do not follow a Gaussian distribution. To address this non-normality, we implemented the Rank-Based Inverse Normal Transformation (RINT) using the Blom formula (alpha = 3.0/8.0) before analysis with the Stepmix. RINT transforms the continuous data into new variables that approximate a standard normal distribution (mean=0, SD=1). This normalization allowed us to appropriately specify the

Gaussian full distribution for the measurement model’s descriptor argument, while the Bernoulli distribution was used for the structural model’s descriptor argument.

As the number of latent classes was not defined *a priori*, we ran with a variable number of classes ranging from 2 to 14. We employed standard model selection criteria, specifically minimizing the Akaike Information Criterion (AIC) or the Bayesian Information Criterion (BIC), to determine the optimal model structure.

Based on this selection process, ten latent classes were determined as suitable. We then ran StepMix using this 10-class model to define the final class membership for all individuals in the cohort. Beyond the latent class model variables, we retrieved additional variables for a more complete phenotypic characterization of the obtained classes, specifically: name data-field instance; centre ID 54; hip circumference 49; BMI 21001; cause death 4000; age at death_40007; age at death_40007; recruitment 53; date of death 40000; ethnicity 21000_i0; height 50; duration of walks 874; moderate activity 894; vigorous activity 914; weight 21002; arterial Stiffness index 21021; liver vol 21080; spleen vol 21083_i2; visceral fat vol 21085; adjusted T/S ratio; 22191; Z-adjusted T/S log 22192; VAT 22407; Trunk-fat vol 22410; abdominal adipose tissue index 22432; weight-to-muscle ratio 22433; abdominal-fat ratio 22434; muscle-fat infiltration 22435; platelet 30080; albumin 30600; alkaline phosphatase 30610; apolipoprotein A 30630; apolipoprotein B 30640; direct bilirubin 30660; urea 30670; calcium 30680; cholesterol 30690; creatinine 30700; C-reactive protein 30710; LDL direct 30780; SHBG 30830; bilirubin 30840; urate 30880.

### Differential Protein Expression (DEP) Analysis

DEP analysis was conducted using the proteomic profiles from Cohort 1a to characterize the signature of each individual risk class against the rest of the cohort (a one-versus-rest comparison).

Statistical testing and correction: For each comparison, the following steps were taken:

- Fold Change (FC): The fold-change of means (FC) was computed to quantify the magnitude of expression difference.
- Significance test: The nonparametric Mann-Whitney U test was employed to statistically compare the protein levels for each protein (using the proteome\_statis script).
- Multiple comparisons correction: The resulting -values were corrected for multiple comparisons using the Benjamini-Hochberg procedure to control the False Discovery Rate (FDR), yielding -values.

Visualization and selection criteria: The statistical significance (q-value) and magnitude of change (log_2_FC) were used to generate volcano plots and select proteins of interest.

· Significance threshold: A stringent threshold of q-value ≤0.001 was applied.
· Upregulated proteins: Selected proteins are marked in red with a log_2_FC ≥ 0.75.
· Down-regulated proteins: Selected proteins are marked in green with a log_2_FC ≤ - 0.75.

Note on visualization: An explicit and strict criterion was applied during plotting to prevent the overlapping of labels on the final visual output, ensuring clarity and readability.

### Adjusted differential protein expression analysis

To refine the understanding of the proteomic signatures, an adjusted DEP analysis was planned. The objective was to determine if the protein level differences identified between the risk classes remained statistically significant after accounting for the potential confounding effects of age, sex, and ethnicity.

Protein selection criteria: The proteins to be included in this adjusted analysis were first selected based on stringent criteria derived from the previous unadjusted DEP results:

· Statistical significance: q-value ≤0.01 (False Discovery Rate).
· Magnitude of Change: log_2_ FC ≥ |0.25|.

To determine the protein level differences between classes after controlling confounders, a Linear Regression analysis was performed for each selected protein.

The analysis utilized the Ordinary Least Squares (OLS) method from the Python statsmodels library (v0.14.4). The model was structured as follows:

- Dependent Variable: Individual Protein Level
- Independent Variables: Class Membership
- Confounding Factors (Adjusted For): Age, Sex, and Ethnicity

This analysis effectively isolates the relationship between class membership and protein level by statistically eliminating the linear effect of the confounding factors.

### Prediction models

Adjusted Cox proportional hazards (Cox PH) Model and protein selection: A separate Cox PH analysis was conducted to evaluate the impact of individual protein levels on survival time. Each model included three covariates: age, sex, and the level of a single protein.

Data preprocessing: Before modelling, each protein level was Z-score normalized using the StandardScaler function from the sklearn (scikit-learn) library. This transformation scaled the data such that the transformed values had a mean of 0 and a standard deviation of 1.

Modelling and replication: The analysis was performed using the survival script and replicated in Cohort 2 in addition to Cohort 1a (implied by the subsequent selection step). Hazard Ratios (HR) from these models were visualized using forest plots.

Protein selection: From the initial set of 33 proteins, a final selection was made based on statistical significance. Proteins were retained only if they had an associated value in both Cohort 1a and Cohort 2. This selection process resulted in 24 proteins. These proteins were characterized as being upregulated in classes 10 and 3 and demonstrated a significant association with survival time across both cohorts.

Logistic regression for outcome prediction: The same 24 selected proteins were subsequently used to assess their individual predictive power for several binary health outcomes: death, T2DM, arterial hypertension, SLD, or liver fibrosis.

Modelling Setup: The prediction was performed using logistic regression models (implemented with the Python scikit-learn library, v1.7.2). Each model included the protein level of interest, along with age, sex, and the first 10 principal components (PCs) of ethnicity as covariates.

Data preprocessing and replication: Prior to building the models, each protein level was Z-score normalized using the StandardScaler from sklearn. This ensured the transformed protein data had a mean of 0 and a standard deviation of 1. The full analysis (including all four outcomes) was performed on cohorts 1a and 2 (except SLD and liver fibrosis in cohort 2).

Performance assessment and visualization: The performance of the linear models was assessed using Receiver Operating Characteristic (ROC) curves. Area under the ROC curves (AUC) values for the 24 proteins across each outcome and cohort were assessed.

### Developing Parsimonious Protein Scores using Lasso Regression

This phase focused on developing parsimonious predictive models for survival using two distinct sets of proteins: the 24 selected proteins and a subset of 20 proteins (where the extra four were LEP, KRT18, AIFM1, and FCAMR). The goal was to create protein scores (weighted sums of protein levels) to predict two binary outcomes:

- Outcome I: Live/Death status at the end of the study.
- Outcome II: Live/Death status 10 years after the study began.

Lasso Model for Feature Selection: The scores were developed using Least Absolute Shrinkage and Selection Operator (Lasso) regression. The Lasso method automatically performs feature selection by tending to set the coefficients of the least important variables to zero, thereby controlling model complexity and identifying the most relevant proteins for prediction.

Model Fitting and Regularization: Normalization, before modelling, all protein levels were Z-score normalized using the StandardScaler from sklearn.

1. Regularization parameter (α) Selection: The optimal regularization parameter, α, which balances model fit and complexity, was determined using the LassoLarsIC function from the scikit-learn library. This function fits the Lasso model using the Least Angle Regression (LARS) algorithm and selects the model that minimizes the Bayesian Information Criterion (BIC).
2. Coefficient calculation: The final coefficients for the optimal models (optimal protein scores) were computed.

Protein scores were calculated independently in both cohorts. We retained the protein scores calculated for cohort 1a for further use because they yielded a much smaller set of proteins (i.e., a more parsimonious model).

### Dynamic Evaluation of Predictive Scores

A robust evaluation of the predictive performance of each protein score and the established FIB-4 index was conducted to assess their ability to predict survival over different time periods.

Survival modelling and performance metric: The core analysis utilized the Cox PH survival analysis function from the Python scikit-survival library (v0.25.0). The predictive performance over time was assessed using a dynamic version of the AUC. The Bootstrap technique was employed to generate the distribution for this dynamic AUC, ensuring a robust measure of uncertainty. Model covariates and evaluation: Both the protein scores and the FIB-4 index were evaluated in models that included sex and age as covariates.

- Protein Scores: Evaluated with age and sex as covariates.
- FIB-4 Index: Defined as a dichotomous variable: 1 if FIB-4 ≥2.67, and 0 otherwise. The overall predictive power of the scores and the FIB-4 index was evaluated.

### ROC Curve Analysis

In addition to the dynamic AUCs, Receiver Operating Characteristic (ROC) curves were calculated for specific time periods. This was performed for the combined protein scores and the FIB-4 index, as well as for the individual proteins that constituted the final scores.

### Survival analysis

The initial step in survival analysis involved defining the event status and follow-up time. The event status (death) was derived from the instance of “age at death” (Data-Field 40007), which determined if an individual experienced the event or was censored (alive at the end of follow-up). The survival time variable was then computed as the number of days between the “date of recruitment” (Data-Field 53) and the end of observation. Specifically, for those who died, survival time was calculated up to the “date of death” (Data-Field 40000). For censored cases, the time was measured up to the study’s cutoff date, 2024-09-01. These two variables (live-death status and survival time) were subsequently utilized for advanced analyses, including Cox survival modelling and mediation analysis. Cox proportional hazards (Cox PH) models were constructed to assess the impact of class membership on survival time. The final model incorporated five variables: death (the event status), survival (follow-up time in days), age, sex, and the previously identified categorical risk classes. The risk classes (class 1 to class 10) were included in the model using one-hot encoding (dummy coding). Class 5 was designated as the reference group (omitted from dummy coding); this structure allows the resulting coefficients for the other classes to directly reflect the relative hazard (risk) compared to the baseline risk established by class 5. The survival time and live-death status variables were designated as the duration and event variables, respectively, for subsequent analysis. To illustrate the clinical impact of the classification, predicted survival curves were generated using the Cox model results. These predictions were calculated using the survival function, fixing age at 60 years and generating separate scenarios for sex across all simulated cases. The survival function was predicted for each risk class by assuming exclusive class membership: the dummy variable for the class being simulated was set to 1, and all other class dummy variables were set to 0. The predicted survival curves for each sex were then plotted, with the X-axis normalized to months, to visually compare the differential risk associated with each proteomic class. The uncertainty of the estimates was quantified by calculating 95% confidence intervals (CI). These intervals were obtained using the bootstrap method with 100 iterations. In addition to the Cox survival modelling performed on the original cohort, this entire statistical survival analysis was replicated independently on the two subcohorts (cohort 1a and cohort 1b).

In addition to assessing the impact of overall class membership on survival time, a stratified Cox PH analysis was performed based on the primary cause of death (Data-field 40001_i0). This analysis focused exclusively on individuals who had experienced the event (death status) and whose primary cause of death belonged to one of the following major ICD-10 Chapters:

- Chapter II: Neoplasms (Cancers)
- Chapters V & VI: Mental and Nervous System Diseases
- Chapter IX: Diseases of the Circulatory System
- Chapter X: Diseases of the Respiratory System
- Chapter XI: Diseases of the Digestive System

Separate Cox PH models were constructed for each selected ICD-10 chapter.

### Pathway analysis

For analysing the over- and underrepresentation of protein pathways, including Reactome and DisGeNet we used the search engine Enrichr (http://amp.pharm.mssm.edu/Enrichr) and Enrichr-KG (https://maayanlab.cloud/enrichr-kg); the p-value is computed using the Fisher exact test, and z is the z-score calculated by assessing the deviation from the expected rank. In addition, we used the FunRich tool to perform functional enrichment analysis on the generated datasets and to create Venn diagrams or bar graphs (http://www.funrich.org). FunRich performs functional enrichment analysis using background databases integrated from heterogeneous genomic and proteomic resources (>1.5 million annotations).

### Causal mediation analysis

To assess mediation, we performed a causal mediation analysis for all-cause mortality (a time-to-event outcome) using Cox PH hazards models. The analytical framework included a binary class indicator as the exposure, a continuous protein level (standardized to Z-scores) as the mediator, and employed bootstrap inference to derive causal effects. We adjusted all models for age and sex a priori. First, we estimated path a (exposure: class → mediator: protein) via linear regression of the protein on the exposure and covariates to obtain α. Next, we estimated the total effect (path c) of the exposure on mortality using a Cox model including the exposure and covariates but excluding the mediator, yielding τ on the log-hazard scale. We then fit a Cox model including the exposure, mediator, and covariates to estimate the direct effect (path c′; τ′) and the mediator’s effect on the outcome (path b; β). The indirect effect was defined as α×β, the direct effect as τ′, and the total effect as τ. Statistical uncertainty for point estimates, standard errors, confidence intervals, and p-values was quantified using nonparametric bootstrapping with resampling with replacement (typically 1000–5000 iterations). We report effects on the log-hazard scale and corresponding hazard ratios, and compute the proportion mediated as (α×β)/τ, focusing on contrasts involving classes 3 and 10.

For binary outcomes (0,1), liver fibrosis, hypertension, SLD and T2DM, in the cohort 1a, we conducted mediator-specific causal analyses to quantify how individual plasma proteins transmit the association between clinical class membership and multiple outcomes, focusing on contrasts involving classes 3 and 10. Protein abundances were standardized to z-scores (mean 0, SD 1) before modelling to improve comparability across markers.

For each protein mediator, we specified a two-model structure: (1) a linear mediator model regressing the z-scored protein on class membership and prespecified covariates (age and sex); and (2) an outcome model using logistic regression (logit link) of each outcome on class membership, the z-scored protein mediator, and the same covariates.

Mediation was framed in a counterfactual decomposition into total, natural direct, and natural indirect (mediated) effects. Effects were estimated on the log-odds scale and reported as B (coefficients) and odds ratios (OR = exp(effect)). For each outcome–protein–class combination, we summarized total, direct, and indirect effects and the proportion mediated. Primary inference emphasized the magnitude and direction of the mediated component.

We evaluated the incremental value of each mediator on two complementary axes: 1-discrimination (AUC): For each outcome–class–mediator combination, we fit paired logistic models on the same analytic sample: a base model (exposure + prespecified covariates) and a mediator-augmented model (base + mediator). Discrimination was quantified using the Area under the ROC curve (AUC). Changes in AUC were summarized descriptively across combinations to characterize the typical shift in discrimination associated with adding the mediator. 2-Parsimony (AIC and ΔAIC) was assessed using the Akaike Information Criterion (AIC), which penalizes model complexity to balance goodness of fit and the number of parameters. We computed ΔAIC = AIC with mediator (total in figures) – AIC base (Direct in figures). Negative ΔAIC indicates that adding the mediator improves the fit–complexity trade-off (i.e., yields a more parsimonious model), whereas positive ΔAIC suggests the opposite. All analyses were implemented in Python (StatsModels) with custom utilities for mediation and diagnostics.

## Extended results

### Mediating role of proteins in intermediate phenotypes

The consolidated summary (**Supplementary Table S12**) demonstrates that specific proteins exert significant mediating effects on the prevalence of MetS components as logistic outcomes for classes 3 and 10. FTCD and FABP1 were the most prominent mediators of association between liver fibrosis and class. For class 3, the mediation was relatively modest: FTCD accounted for an OR (odds ratio) of ∼1.38 (proportion mediated ∼ 20.8%) with a total OR of ∼3.46, and FABP1 accounted for an OR of ∼ 1.19 (∼ 11.5%) with a total OR of ∼2.97. However, in the high-risk class 10, the proteins accounted for a far greater proportion of the large total ORs (6-7). FTCD alone mediated an indirect OR of ∼ 2.63, representing (∼ 52.9%), with FABP1 contributing an additional indirect OR∼ 1.79 (∼ 30.9%). ADGRG1 provided a moderate mediated effect (indirect OR ∼ 1.50; ∼ 22.7%) in Class 10, while GAST mediation was consistently low (∼ 0.5–6% across classes).

FTCD and FABP1 were the primary mediators of the association between the exposure classes and hypertension. For class 3, FTCD and FABP1 mediated indirect ORs of ∼1.30 (∼19.3%) and ∼1.20 (∼14.2%), respectively, contributing to a total OR of ∼3.7–4.0. In class 10, FTCD (indirect OR ∼ 1.85; ∼ 17.4%) and ADGRG1 (indirect OR ∼ 1.56) accounted for similar mediated proportions, at ∼17.4% and ∼15.8%, respectively. GAST’s contribution was negligible, consistently below 2%. The protein-mediated effects for severe liver disease (SLD) were the most substantial signals found in the analysis. Across both classes, FTCD was the primary mediator, though the pattern of contribution differed between the groups. In class 3, FTCD’s indirect OR ∼ 1.70 (∼ 21.3%) and FABP1’s OR of∼ 1.31 (∼ 11.0%) paralleled very large total ORs ∼ 11–12. In class 10, FTCD dominated (OR of∼ 3.40; ∼ 34.0%), followed by ADGRG1 (OR of∼ 2.00; ∼ 20.5%) and FABP1 (OR of∼ 1.64; ∼ 14.8%), consistent with total ORs ∼ 29–37.

For the T2DM outcome, FTCD and ADGRG1 were the leading mediators. For class 3, FTCD’s indirect OR was ∼1.28 (∼31.6%), with a total OR of ∼2.20; ADGRG1 (OR of ∼1.13; ∼12.8%) and FABP1 (OR of ∼1.11; ∼11.7%) contributed moderately. For class 10, FTCD (OR of ∼ 1.73; ∼31.3%) and ADGRG1 (OR of∼ 1.60; ∼25.6%) aligned with large total ORs of ∼ 5.7–6.5, while GAST and FABP1 showed smaller but consistent mediation (OR of ∼ 1.16 and ∼ 1.19; ∼ 8–9%, respectively). Full details are presented in **Supplementary Table S12**.

Furthermore, **Supplementary Fig S6** shows the gains in AUC and AIC goodness-of-fit from including the mediator in the model for each outcome.

## SUpplementary Figures

**Supplementary Figure S1:**
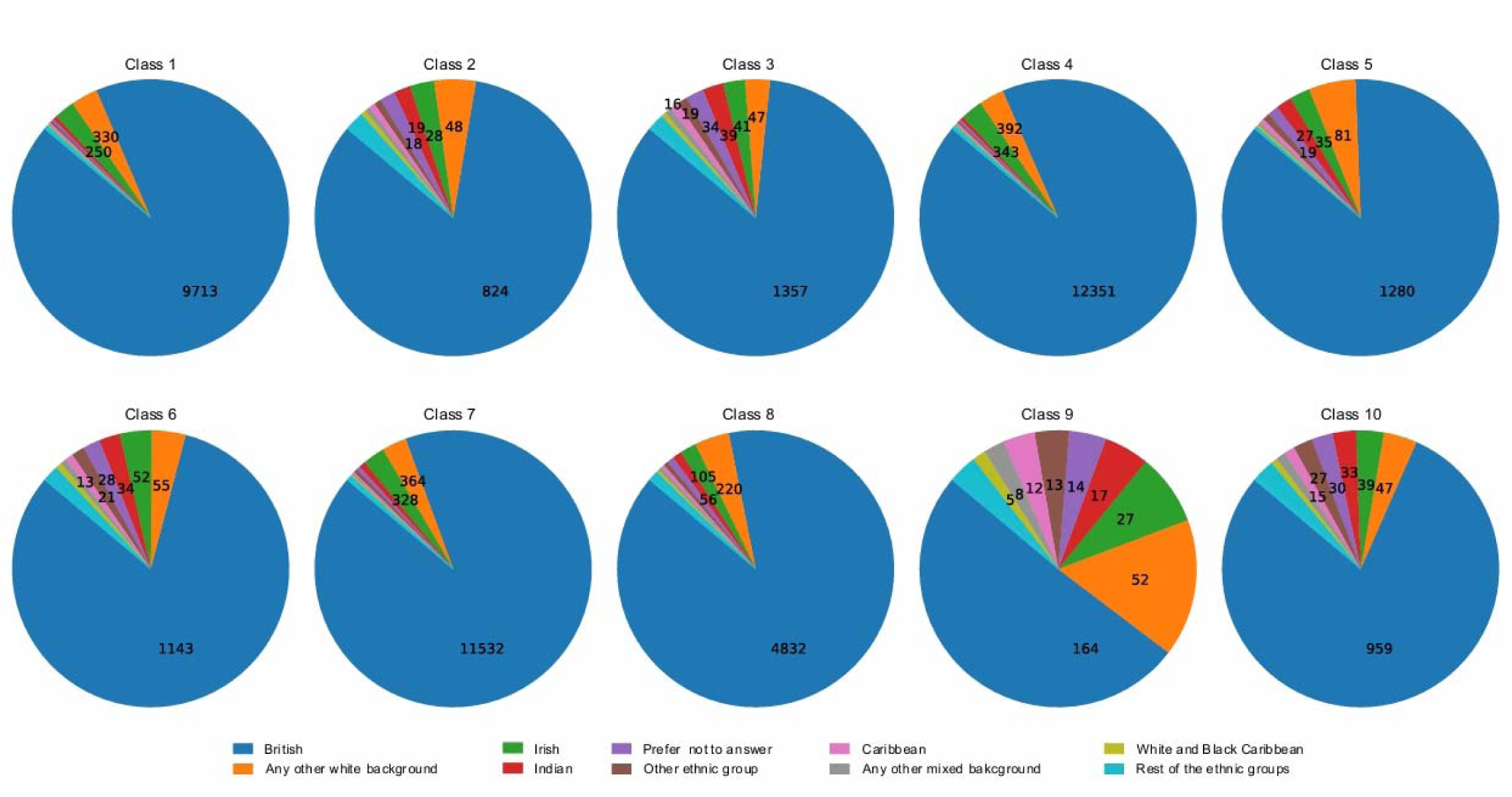
**Participant ethnicity distribution by class in cohort 1**

**Supplementary Figure S2:**
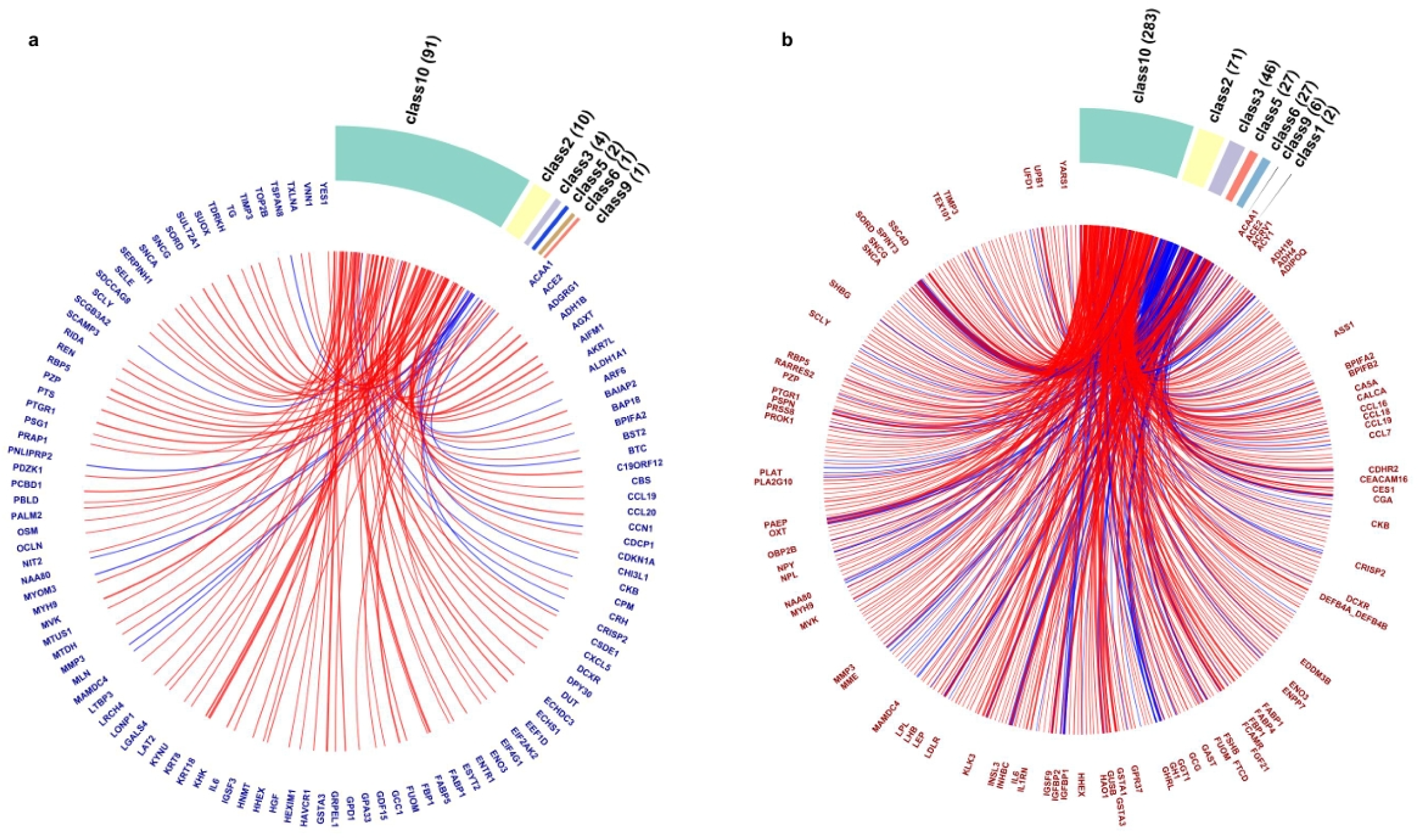
**Proteomic profiles across latent classes.**Circos plots showing the distribution of proteins across latent classes. Connections (edges) represent proteins linked to their class of origin, where red edges denote upregulated proteins and blue edges denote downregulated proteins relative to the other classes drawn proportionally to fold-change magnitude. **a**: proteins uniquely detected in each class, filtered by a cutoff of log2 Fold Change (FC) >=|0.58| (∼FC: 1.79 for upregulated and ∼0.6 for downregulated proteins, respectively) to highlight class-specific molecular signatures. **b**: proteins shared among classes, displayed using a cutoff of log2 FC >=| 0.4| (∼FC: 1.5 for upregulated and ∼0.7 for downregulated proteins, respectively) to emphasize abundance differences. In both cases, cutoffs were applied for the sake of readability. Circos plots were generated in R using the *circlize* package.

**Supplementary Figure S3:**
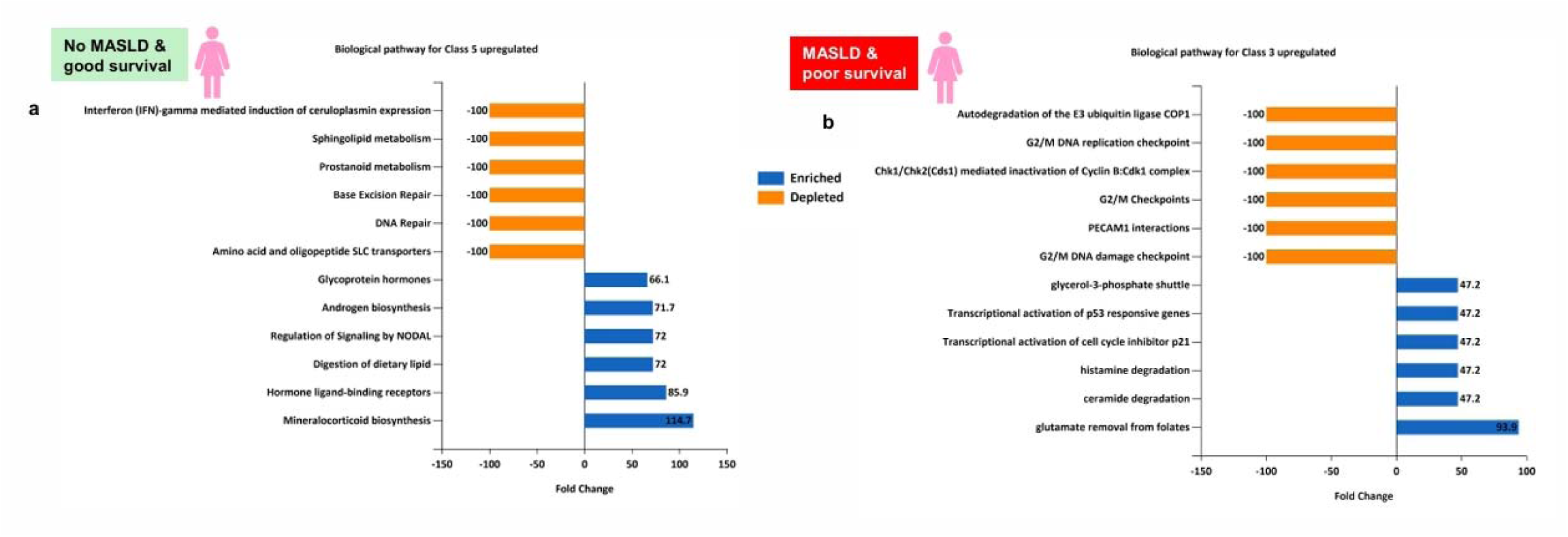
**Divergent protein profile according to survival among females with and without MASLD.** Enrichment of biological pathways associated with poor or good survival in classes with and without MASLD.

**Supplementary Figure S4:**
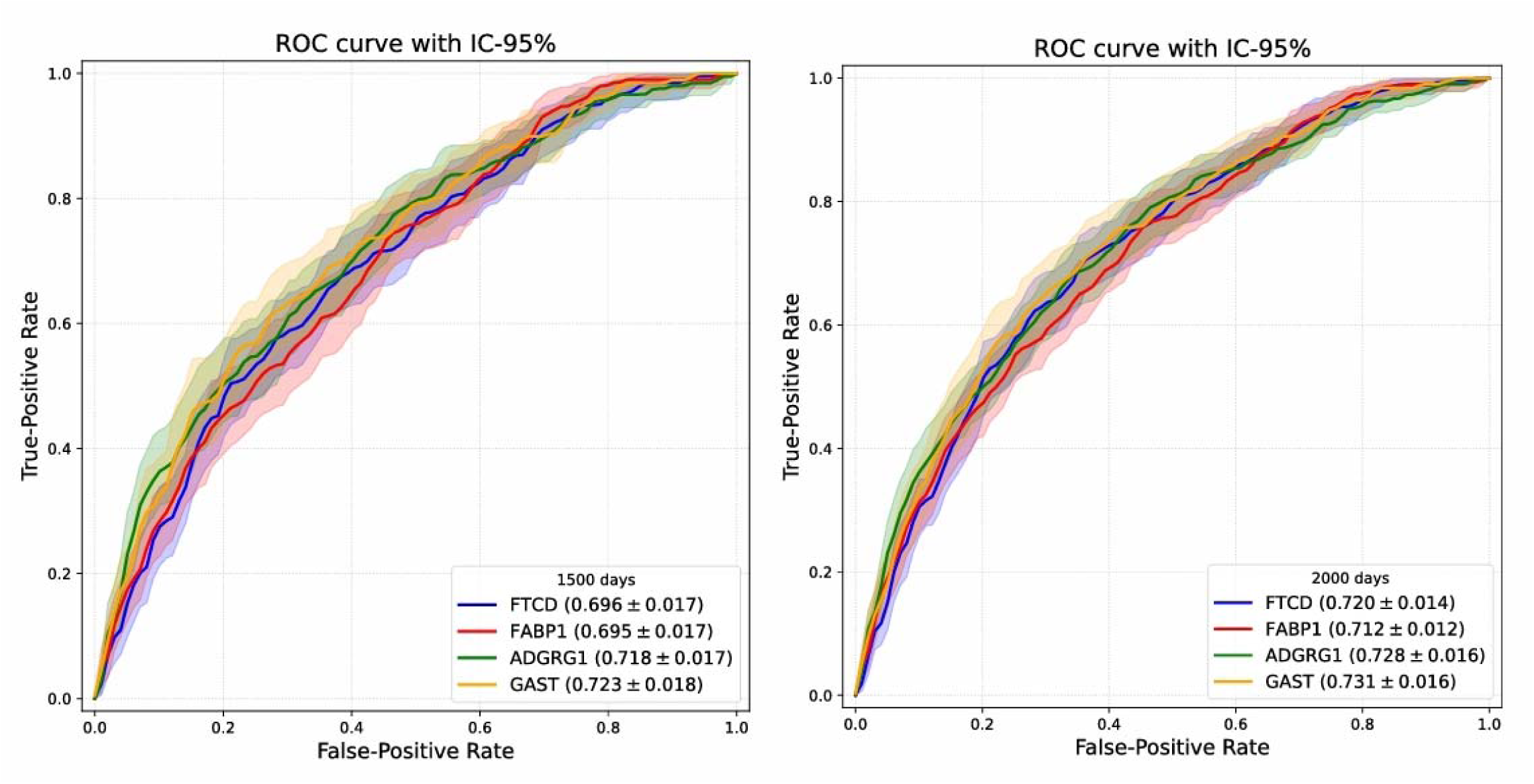
**Performance of the PS4 proteins in predicting all-cause mortality over different times.** ROC curves for FTCD, FABP1, ADGRG1, and GAST for predicting all-cause deaths at 1500 and 2000 days of follow-up adjusted by age, sex, and ethnicity. Shadow bands stand for 95% confidence Intervals. Results are expressed as AUC±SE adjusted by age and sex.

**Supplementary Figure S5:**
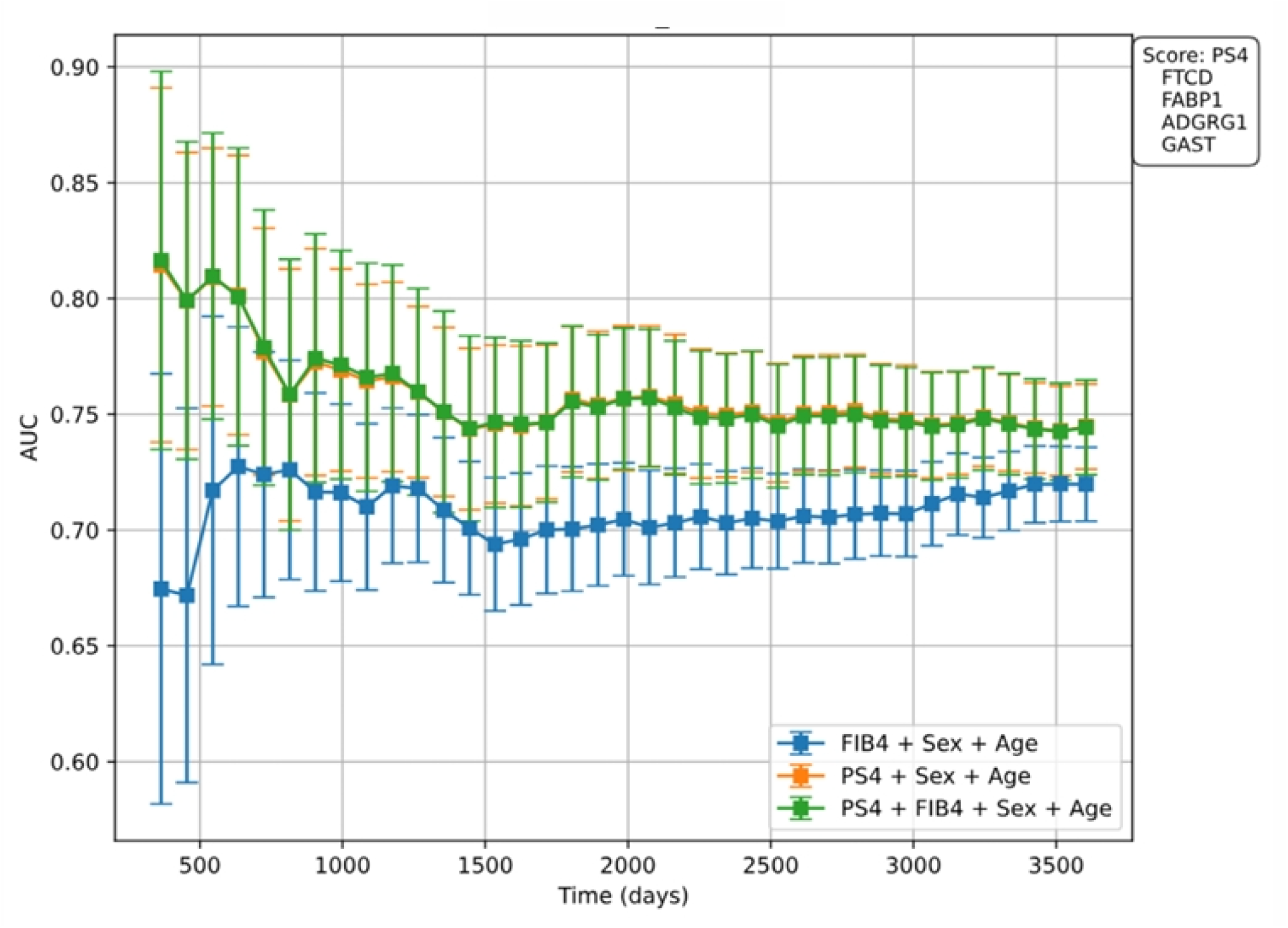
Protein set replicated externally in Icelanders. Discrimination power of the different models described in the legend panel for death during the follow-up period (in days). AUCs for all individuals of cohort 2, N = 44,252. Results are shown as AUC±95 % CI at each follow-up time point in days (death within 1500-2000 days). PS4 stands for the protein score obtained by LASSO with BIC, using the remaining 20 proteins after subtracting the four proteins not replicated in the Iceland cohort (LEP, KRT18, AIFM1, and FCAMR) as described in the main text. FIB-4 stands for fibrosis-4 index, a predictor of liver fibrosis.

**Supplementary Figure S6:**
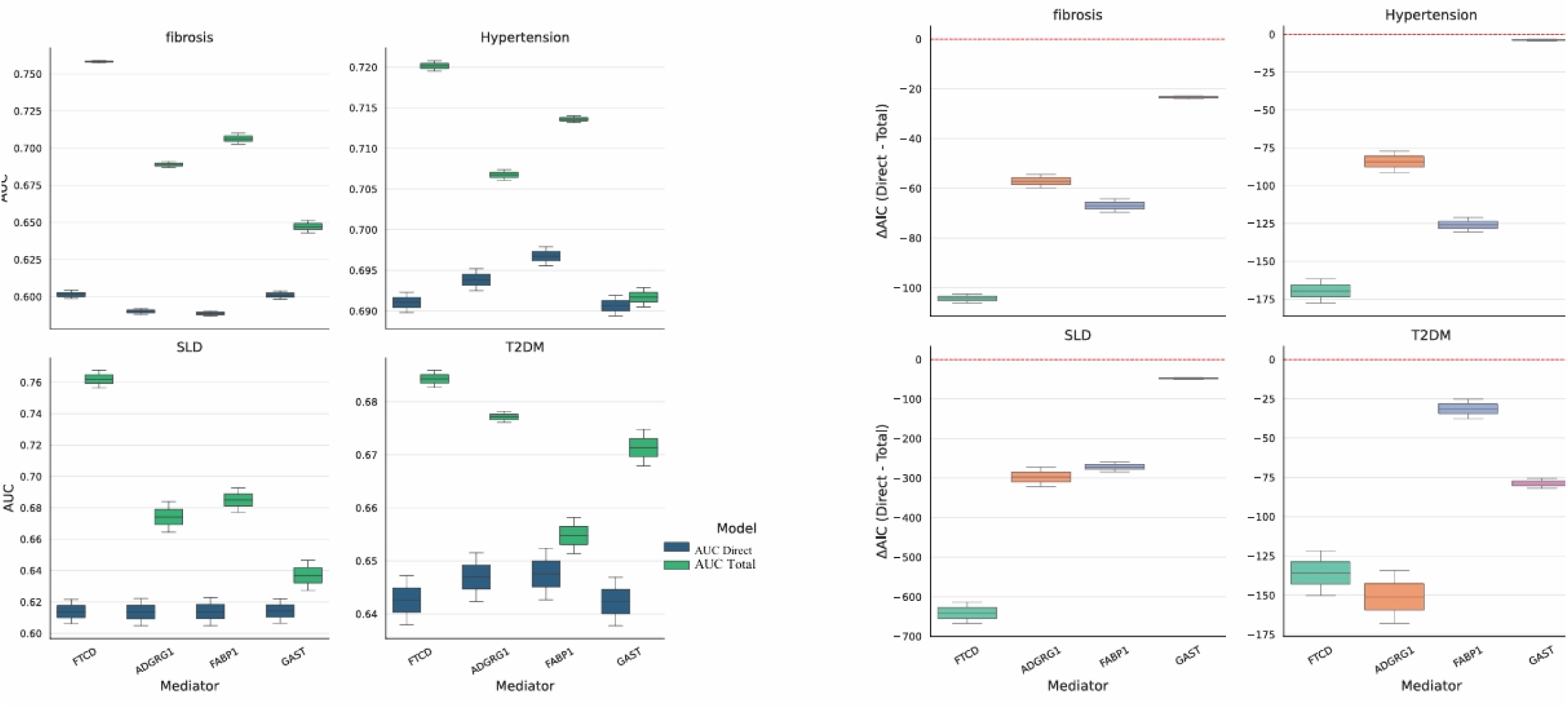
Diagnostic performance by mediators. **a**: (AUC distributions; base vs mediator-augmented). For each mediator (FTCD, ADGRG1, FABP1, GAST), paired boxplots summarize the distribution of AUC values for the base model (without mediator, “Direct”, blue boxes) and the mediator-augmented model (‘total”, green boxes), aggregating across outcomes and classes. Upward shifts in AUC from the base to the augmented model reflect improved discrimination when the mediator is included. Even modest median gains can be meaningful if accompanied by consistently negative ΔAIC, suggesting that the mediator improves both discrimination and the fit-complexity balance. AUCs were computed on matched complete-case samples. Because AUC changes can be small in logistic settings with strong baseline predictors, joint consideration with ΔAIC provides a more nuanced view of incremental value. **b**: (ΔAIC distributions; with base, direct – mediator included total model). Boxplots depict the distribution of ΔAIC values (AIC direct– AIC total, with mediator) for each mediator, aggregating across outcomes and classes. Boxes show IQRs, central lines indicate medians, whiskers extend to 1.5×IQR. Negative medians (and a predominance of negative values) indicate that including the mediator yields more parsimonious models, i.e., the improvement in goodness-of-fit outweighs the penalty for added parameters. The spread reflects heterogeneity of the mediator’s value across outcome–class combinations. ΔAIC values were computed on the same analytic samples used for AUC. Results are intended as complementary to mediation effect estimates rather than as formal hypothesis tests.

## Supplementary tables

https://doi.org/10.5281/zenodo.17764543

**Supplementary Table S1: Quantitative analysis of latent classes in MASLD**. Cohort 1. Clinical, anthropometric, and biochemical variables stratified by sex.

**Supplementary Table S2: Cohort 1a.** Clinical, anthropometric, and biochemical variables stratified by sex.

**Supplementary Table S3: Full list of Olink™ Explore 3072 protein analytes and targeted panels.** This table lists the 2,923 unique protein analytes measured using the Olink™ Explore 3072 Proximity Extension Assay (PEA). The analytes are captured across eight panels: Cardiometabolic, Cardiometabolic II, Inflammation, Inflammation II, Neurology, Neurology II, Oncology, and Oncology II.

**Supplementary Table S4: Proteomic profiles in MASLD subgroups.** Complete set of differentially expressed proteins in each latent class.

**Supplementary Table S5: Pathway analysis of differentially expressed proteins in MASLD subgroups.** Examination of the diseases prioritised by the database DisGeNET (https://www.disgenet.com).

**Supplementary Table S6: Cohort 1b.** Clinical, anthropometric, and biochemical variables stratified by sex.

**Supplementary Table S7: Cohort 2.** Clinical, anthropometric, and biochemical variables stratified by sex.

**Supplementary Table S8:** The 24-set of proteins selected to model a protein score for predicting all-cause deaths.

**Supplementary Table S9:** Results of cross-library analysis of 24-protein signature using Enrich-KG (https://maayanlab.cloud/enrichr-kg).

**Supplementary Table S10: Proteins Replicated in the deCODE Icelandic Cohort.** This table lists the protein set that was successfully replicated in an external population sample from the deCODE dataset (Eiriksdottir T, Ardal S, Jonsson BA, Lund SH, Ivarsdottir EV, Norland K, et al. Predicting the probability of death using proteomics. Commun Biol 2021;4:758), which includes data from the Icelandic cancer project.

**Supplementary Table S11: Mediation analysis of proteomic signatures on mortality** Decomposition of the total effect and mediation role for PS4.

**Supplementary Table S12: Mediating role of proteins in intermediate phenotypes.** Decomposition of the total effect and mediation role for PS4 for liver fibrosis, steatotic liver disease, arterial hypertension, and type 2 diabetes.

